# Identification of *de novo* variants from parent-proband duos via long-read sequencing

**DOI:** 10.1101/2025.02.24.25322424

**Authors:** Leandros Boukas, Emmanuèle C. Délot, Georgia Pitsava, Christine Lambert, Cairbre Fanslow, Primo Baybayan, Sami Belhadj, Bojan Losic, John Harting, Krista Bluske, Jonathan LoTempio, Huda B. Al-Kouatly, Rachid Karam, William J. Rowell, Changrui Xiao, Eric Vilain, Seth I. Berger

## Abstract

While *de novo* variants cause many Mendelian disorders, their detection currently requires sequencing of the proband and both biological parents. This is not feasible when only one parent is available, a limitation for millions of families. Here, we develop *duoNovo*, which identifies *de novo* variants from parent-proband duos using long-read sequencing followed by haplotype reconstruction and detection of identical-by-descent haplotype blocks. We sequenced 117 trios with PacBio HiFi sequencing and applied *duoNovo* to each of the 234 duos constructed by masking one parent, classifying over 60 million variants according to their *de novo* status. We evaluated *duoNovo*’s performance against classifications obtained using the full trios (which included over 10,000 *de novo* variants), demonstrating very high precision (∼ 98% among variants absent from gnomAD) and low error rate. In a cohort of 74 duos undergoing diagnostic evaluation, *duoNovo* prioritized a *de novo* intronic variant likely to disrupt splicing, while also uncovering the *de novo* status of two variants already classified as pathogenic. In summary, *duoNovo* has the potential to significantly increase the diagnostic yield of single-parent genetic testing, and represents an example where long-read sequencing provides clear benefit over short-read sequencing even for single nucleotide variants. It is freely available as an R package (https://github.com/sbergercnmc/duonovo).

## Introduction

*De novo* variants, which are present in affected probands but not (constitutively) present in the parents, underlie a substantial fraction of Mendelian disorders (Brunet et al., 2021; Ligt et al., 2012; Posey et al., 2017; Vissers et al., 2010; Wright et al., 2023; Yang et al., 2014). These disorders are typically highly penetrant, and as a result, the American College of Medical Genetics (ACMG) guidelines use a variant’s *de novo* status as a strong criterion for pathogenicity (Richards et al., 2015).

Currently, *de novo* status can only be determined through sequencing of the proband and both biological parents. However, various factors can prevent both parents from participating in genetic testing. US Census statistics indicate that there are approximately 10 million single-parent families in the United States alone (predominantly single-mother families). Additional reasons that make it infeasible to obtain samples from both biological parents include active military duty, children conceived through sperm or egg donation, or parental death. Consequently, probands in these families face a higher risk of non-diagnostic results (Rehm et al., 2023; Richards et al., 2015; Wojcik et al., 2024). Given that ACMG guidelines advise against using VUS in treatment plan determinations, this can lead to missed opportunities to benefit from precision treatment and management options. Moreover, the magnitude of the reduction in diagnostic yield due to the unavailability of parental samples correlates with ancestry, indicating the potential for further exacerbation of existing health disparities (Wright et al., 2023).

Long-read sequencing (LRS) is emerging in the clinical setting, although higher cost combined with uncertainty regarding diagnostic benefit over short-read sequencing (SRS) currently limit its widespread adoption (Wojcik et al., 2023). To characterize the potential diagnostic benefit, a large body of work has focused on the ability of LRS to detect large/complex structural variants and other variants missed by SRS (Cohen et al., 2022; Dominguez Gonzalez et al., 2025; Mastrorosa et al., 2023; Merker et al., 2018; Miller et al., 2021; Mizuguchi et al., 2021; Negi et al., 2025; Xie et al., 2020); an additional studied benefit is the concurrent detection of epigenetic variation (Vollger et al., 2025). However, recent improvements in LRS technologies, such as the development of PacBio HiFi LRS, now allow one to obtain reads with very high basecall accuracy (Gustafson et al., 2024; Hiatt et al., 2024; Kolmogorov et al., 2023; Kucuk et al., 2023; Mastrorosa et al., 2023; Wenger et al., 2019), enabling the identification of small variants including single-nucleotide variants. One unique advantage of LRS is that it couples the identification of such variants with their reliable, read-backed phasing (Holt et al., 2024; Martin et al., 2016). This is especially important for potentially disease-causing variants evaluated for *de novo* status in the clinical diagnostic setting, which are very rare or even unique to an affected individual and thus not amenable to phasing methods that rely on information about haplotype frequencies in the population (Browning and Browning, 2011; Guo et al., 2024).

Here, we leverage these features of LRS to develop a method - which we call *duoNovo* - to identify *de novo* variants from duos (proband and one biological parent), with the potential to open more diagnostic opportunities for these families.

## Methods

### Participants

We sequenced 117 trios. 22 of these trios were derived from 11 families with 2 siblings each, which were each split into two trios, whereas 6 trios were derived from two families with 3 siblings each, which were each split into 3 trios. Age (at childbirth) and genetic ancestry of the parents are provided in Supplemental Table 1.

### Long-read sequencing

Samples were prepared for LRS following the standard operating procedure (SOP) available at PACB.com. (”Preparing whole genome and metagenome libraries using SMRTbell prep kit 3.0”). Sequencing was performed on the PacBio Revio system with the Revio polymerase Kit, following the Revio SMRT Link setup. Genomic DNA quality and concentration were assessed using the FEMTO Pulse (Agilent Technologies) and Qubit dsDNA HS reagents Assay kit (Thermo Fisher Scientific). SMRTbell libraries were constructed on the Hamilton Microlab Star and VENUS 5 system following the SOP available on PACB.com under ”Preparing whole genome and metagenome libraries using SMRTbell prep kit 3.0”. For library characterization, post size-selected SMRTbell Library samples were quantified using the Qubit DNA HS assay and DNA size was estimated using the FEMTO Pulse. The libraries were loaded at an On-Plate Concentration of 90 pM.

Raw sequencing reads were subsequently processed as follows. Circular consensus sequences were generated from subreads using CCS (version 7.0.0) with default settings, yielding demultiplexed HiFi reads. These reads were then aligned to the hg38 reference genome (https://github.com/PacificBiosciences/reference_genomes/blob/main/reference_genomes/human_GRCh38_ no_alt_analysis_set/README.md) using PBMM2 (version 1.10.0), with default parameters.

### Variant calling

Variant calling following LRS was performed using DeepVariant (version 1.5.0; (Poplin et al., 2018; Yun et al., 2021)). For each duo as well as for each trio, gvcfs were first generated and joint variant calls were subsequently produced for each duo and trio separately with GLnexus (version 1.4.1) with the preset “DeepVariant unfiltered”. All our analyses and numbers reported are based on candidate *de novo* variants that were called both from a duo and its corresponding trio and thus were candidates we could evaluate.

For the evaluation of *duoNovo*’s performance, *de novo* classifications were determined to be correct if the missing parent did not have the candidate variant allele at that position (that is, if the non-sequenced parent was either homozygous for a non-variant allele or heterozygous with two different non-variant alleles). Similarly, non-*de novo* classifications were determined to be correct if the non-sequenced parent had the candidate variant allele at that position.

To obtain *de novo* variants from the entire trio, we used DNM2 (naive model) to identify sites with a violation of Mendelian inheritance indicative of a *de novo* event (heterozygous variant in the proband; homozygous reference call in both parents). Out of these sites, we then retained those with a minimum GQ of 30 for each sample, a minimum depth of 20 reads for each sample, and 0 reads supporting the alternate allele. Multi-allelic variant sites were excluded. Finally, during our analysis we discovered 13 trios with many artifactual *de novo* calls from DNM2 due to the fact that variants from either one or both parents were called using a different version of the PBMM2 aligner (1.16.99) than the version used for the proband (1.10.0); these trios were excluded from our sensitivity analyses.

### Phasing

Following LRS, phasing was conducted using HiPhase (version 1.4.0) (Holt et al., 2024) with default parameters on the joint called vcf from each duo. HiPhase assigned each phased variant to a phasing set. Variants in the same phasing set had the same phase relative to one another, enabling the resolution of those variants into haplotypes.

### Estimation of haplotype similarity and classification of candidate *de novo* variants with ***duoNovo***

Following phasing, we imported the vcf files containing the phased variant calls into R with the VariantAnnotation package (Obenchain et al., 2014). We then only retained variant positions where sequencing depth was at least 20, and GQ was at least 30. As stated in the Results, we defined candidate *de novo* variants as variants heterozygous in the proband and absent in the parent.

We then developed *duoNovo* to obtain classifications for these candidate variants. *duoNovo* first defines haplotype blocks as genomic regions within which: a) proband variants are assigned to the same phasing set; and b) parent variants are assigned to the same phasing set. *duoNovo* then evaluates candidate variants, separately in each of the two phasing orientations. It starts by discarding candidate variants that are either in haplotype blocks smaller than 10 kb, or at the boundaries of the remaining blocks (2 kb from the start/end coordinates). These boundary variants are discarded because we found that when they are classified as *de novo*, they tend to be false positives, especially in mother-proband duos (see below). Subsequently, *duoNovo* sequentially examines each of the proband haplotype blocks containing candidate *de novo* variants. Each of these haplotype blocks is compared to each of the two corresponding parental haplotype blocks using the Hamming distance. In addition, *duoNovo* performs the same comparisons for the other proband haplotype (not containing the candidate variant). In all cases, the Hamming distance between a pair of haplotype blocks is calculated after representing each haplotype block as a binary string (with 0 for the reference allele and 1 for the variant allele) and excluding candidate variants in the same phasing orientation as the candidate variant being evaluated. After performing these comparisons, *duoNovo* produces classifications based on the following criteria:

(i) If the proband haplotype block containing the candidate variant has a Hamming distance of 0 with only one of the two parental haplotype blocks, while the other proband haplotype block has a Hamming distance greater than 40 with both parental haplotype blocks, the candidate variant is classified as *de novo* (see section “Parameter Sensitivity Analysis” for an examination of the impact of the specific Hamming distance threshold).
(ii) If the proband haplotype block containing the candidate variant has a Hamming distance greater than 40 with both parental haplotype blocks, while the other proband haplotype block has a Hamming distance of 0 with only one of the two parental haplotype blocks, the candidate variant is classified as inherited from the missing parent.
(iii) If none of the two above conditions were satisfied (e.g. due to no proband-parent haplotype pair having a Hamming distance of 0), the candidate variant does not receive a classification and is labeled as uncertain.

To minimize false positive classifications due to genotyping errors, from the resulting classifications we exclude variants falling within regions stratified as problematic by the Genome-in-a-Bottle consortium (Dwarshuis et al., 2024).

Finally, if two or more of the variants classified as *de novo* are present within the same haplotype block, *duoNovo* discards all *de novo* classifications in that block and labels these variants as present on a “multi-*de novo* haplotype”. While these could represent true *de novo* events, we have observed that they tend to be false positives (see section “Examining sources of false positive classifications”).

### Interpretation of Hamming distance equal to 0 between a proband-parent haplotype block pair

Intuitively, a Hamming distance equal to 0 between a proband-parent haplotype block pair indicates that this haplotype is shared identical-by-descent. This intuition can be made more precise when we consider the genotypes of the different positions that determine the Hamming distance.

1. **Positions where the proband is heterozygous and the parent is homozygous.** These positions provide direct information about haplotype transmission, because only one of the two proband haplotypes could have been transmitted from the sequenced parent. We note that, without read-backed phasing, candidate variants would always be classified as inherited from the non-sequenced parent, since they are absent in the sequenced parent. This illustrates the critical role of read-backed phasing. In turn, this implies that in order for the classification to be accurate, it is critical to ensure that the read-backed phasing of variants in the region is accurate. We accomplish this using the two types of positions below.
2. **Positions where both proband and parent are heterozygous**. When considered in isolation, these positions do not provide direct information about haplotype transmission, because each of the two proband haplotypes could be identical-by-descent with the corresponding parental haplotype. However, when considered in the context of the haplotype inferred to have been transmitted from the sequenced parent based on 1 above, these positions serve as a quality control for the accuracy of the phasing in the region, in both the proband and the parent.
3. Positions where the proband is homozygous and the parent is heterozygous**. Like 2, these**

do not provide direct information about haplotype transmission in isolation, because each of the two proband haplotypes could be identical-by-descent with the parental haplotype that harbors the allele for which the proband is homozygous. However, when considered in the context of the haplotype inferred to have been transmitted from the sequenced parent based on 1 above, they serve as a quality control for the accuracy of phasing in the parent, thus lending further support to the accuracy of phasing in the proband as described in 2.

Consequently, when the Hamming distance between a pair of proband-parent haplotype blocks is equal to 0, we can infer that: a) the proband inherited this haplotype block from the sequenced parent, and not from the other (non-sequenced) parent; b) the phasing of the candidate variant based on which we have determined whether the variant is *de novo* or not is accurate.

### Variants that fail QC versus variants that are classified as uncertain

The following candidate variants are excluded by *duoNovo*’s QC filters:

1. Variants that do not pass sequencing depth and/or GQ thresholds. These thresholds are tunable parameters.
2. Variants that cannot be resolved into a haplotype block because HiPhase did not assign them to a phasing set (in either the proband of the parent).
3. Variants in haplotype blocks whose size is smaller than the minimum chosen threshold. This threshold is a tunable parameter.
4. Variants too close to the boundaries of a haplotype block, based on the chosen threshold for the distance from the boundary. This threshold is a tunable parameter.
5. Variants within problematic regions (for example, as annotated by Genome-in-a-Bottle). One can choose whether or not to supply a list of such problematic regions.

The first 4 categories are variants for which *duoNovo* does not generate a classification. The final category (variants in problematic regions) are labeled as variants that failed QC, but we also output the classification that would have been received, had they not been within a problematic region.

By contrast, candidate variants that get classified as uncertain are variants that passed all the above QC filters, but the corresponding haplotype pair comparisons (based on the chosen Hamming distance threshold, which is a tunable parameter) did not satisfy the criteria required to receive a classification as either *de novo* or present on the missing parent’s haplotype.

### Variant classification using proband-sibling-parent trios

For our analysis of variants using proband-sibling-mother and proband-sibling-father trios, we first constructed these trios from the families with siblings. Each sibling pair was analyzed twice, alternating which sibling was designated as the proband. This is because the set of candidate variants that are evaluated for *de novo* status is proband-specific (being derived from the corresponding proband-parent duo). There were two families with 3 siblings each, from which 3 different proband-sibling-mother and 3 different proband-sibling-father trios were constructed.

Using these proband-sibling-parent trios, we generated joint called phased trio vcf files in an identical fashion as the regular proband-father-mother joint called phased trio vcfs.

For variant classification, we first obtained haplotype blocks within which: a) proband variants are assigned to the same phasing set; b) sibling variants are assigned to the same phasing set; c) parent variants are assigned to the same phasing set. We subsequently defined candidate variants (heterozygous variant call in the proband; homozygous reference call in both the sibling and the parent), and proceeded to classify them. Specifically, a candidate variant was classified as *de novo* on the missing parent’s haplotype, if all of the following were satisfied:

(i) The proband haplotype block containing the candidate variant had a Hamming distance of 0 with exactly one of the two sibling haplotype blocks.
(ii) The proband haplotype block without the candidate variant had a Hamming distance of 0 with exactly one of the two parental haplotype blocks.
(iii) The proband haplotype block containing the candidate variant had a Hamming distance greater than 40 with both parental haplotype blocks.
(iv) The proband haplotype block without the candidate variant had a Hamming distance greater than 40 with both sibling haplotype blocks.

When assessing the gain in sensitivity by using siblings as surrogates for the missing parent (Figure 4A), we obtained candidate variants among those previously classified – from the corresponding duo – as either present on the haplotype inherited from the missing parent, or uncertain. When assessing the genome-scale positive predictive value of the sibling trio-based approach (Figure 4B), candidate variants were obtained from the entire genome regardless of their previous classification from the corresponding duo.

### Parameter sensitivity analysis

We chose the first 80 duos sequenced (40 father-proband; 40 mother-proband) to examine the impact of *duoNovo*’s tuning parameters on the positive predictive value (PPV) and the number of *de novo* classifications.

First, we varied the threshold for the Hamming distance used to determine if a pair of proband-parent haplotype blocks is dissimilar and is thus not shared identical-by-descent (our default being 40). We found that a threshold of 0 always yields more *de novo* classifications compared to our default (as expected, since it is more lenient), but almost always yields a lower PPV, indicating that these classifications are enriched for false positives (Supplemental Figure S10, S11). Generally, we observed slight improvement when increasing thresholds up to 40, but no net gain when using thresholds above 40.

Second, we varied the distance threshold from the boundaries of haplotype blocks. By default, *duoNovo* excludes variants within 2kb of the start/end coordinates of haplotype blocks. We found that using a more lenient threshold tends to yield more false positives, especially in mother-proband duos (potentially reflecting a drop in phasing accuracy at these boundaries; Supplemental Figure S12). The impact on the number of *de novo* classifications is minimal, as expected given that this threshold affects the inclusion of a comparatively small number of variants (Supplemental Figure S13).

We then varied the thresholds for GQ and sequencing depth. One would naturally expect that using more lenient thresholds for candidate variants themselves would lead to an inflation of false positives. However, the impact of these thresholds when focusing on the positions surrounding candidate variants – which determine the Hamming distance between each proband-parent haplotype block pair – is less obvious. We found that, while using more lenient GQ thresholds (10 or 20 instead of 30) appears to increase the PPV (Supplemental Figure S14), this is because a much smaller number of variants get classified as *de novo*. This is likely due to the fact that a lower GQ causes haplotype block pairs that in reality are identical-by-descent to have a Hamming distance greater than 0 due to sequencing errors, thus leading to missed *de novo* variants. Finally, a more lenient sequencing depth threshold (10 instead of 20) had an overall small impact (Supplemental Figure S15).

### Targeted variant analysis

For targeted variant analysis (that is, when testing variants for *de novo* status after manual curation has already deemed them to be of interest), we consider it reasonable to use more relaxed thresholds for the different *duoNovo* parameters. This is because these variants have a higher prior probability of being *de novo*.

For our targeted analysis of variants from the UCI-GREGoR case set, we used a sequencing depth cutoff of 10 and a GQ cutoff of 20. Subsequently, a candidate variant was:

- Classified as *de novo* if the proband haplotype block containing the variant had a Hamming distance ≤ 2 with only one of the two parental haplotype blocks (and a Hamming distance > 2 with the other parental haplotype block), while the other proband haplotype block had a Hamming distance > 5 with both parental haplotype blocks.
- Classified as present on the missing parent’s haplotype if the proband haplotype block containing the variant had a Hamming distance greater than 5 with both parental haplotype blocks, while the other proband haplotype block had a Hamming distance ≤ 2 with only one of the two parental haplotype blocks (and a Hamming distance > 2 with the other parental haplotype block).

For this targeted variant analysis we did not apply any filter for the distance of the variants to the haplotype block boundary.

### Examining sources of false positive classifications

As described above, from the resulting classifications *duoNovo* excludes *de novo* variants clustered in the same haplotype block, and variants within regions annotated as problematic by the Genome-in-a-Bottle consortium (Dwarshuis et al., 2024). This is because we found that including these variants leads to an increased rate of false positive *de novo* classifications. Specifically, after pooling counts across all father-proband duos, we found that including variants within Genome-in-a-Bottle problematic regions reduces the positive predictive value from 91% to 79% in father-proband duos, and from 72% to 61% in mother-proband duos. Including variants classified as *de novo* and clustered in the same haplotype block reduces the positive predictive value to 68% in father-proband duos, and to 46% in mother-proband duos.

Finally, we found that both of these sources of false positive classifications have a smaller impact when restricting to variants absent from gnomAD. In that case, including variants within Genome-in-a-Bottle problematic regions reduces the aggregate positive predictive value to 96% in father-proband duos, and to 86% in mother-proband duos. Including variants classified as *de novo* that are clustered in the same haplotype block reduces the aggregate positive predictive value to 98% in father-proband duos, and to 93% in mother-proband duos.

### Variant Annotation

We annotated all variants using ANNOVAR (Wang et al., 2010). For each variant, annotations included genomic compartments (e.g. exonic, intronic, intergenic), CpG vs non-CpG context, gnomAD version 4.1 allele frequencies, and whether the variant falls into a GIAB-problematic region (Dwarshuis et al., 2024).

### Relatedness and genetic ancestry calculation

Across all pairs of samples, relatedness was calculated using the “relate” function from Somalier (Pedersen et al., 2020) with default parameters. Genetic ancestry for each sample was predicted using the Somalier “ancestry” function using labeled data from the 1000 genomes project.

### Mutation types

We calculated the number of occurrences of the different types of single nucleotide variants per duo with the MutationalPatterns R package (Manders et al., 2022). For Supplemental Figure S4, percentages were calculated after pooling the counts across all father-proband duos, and all mother-proband duos. The mutation types tested for a different probability of occurrence in the paternal vs the maternal germline were C>A, C>G, C>T within and outside of the CpG context, T>A, T>C, T>G.

### *De novo* variant curation from UCI-GREGoR duos

Variants classified as *de novo* from the UCI-GREGoR case set duos were annotated with ANNOVAR (as described above) and filtered to only retain variants that were: i) absent from gnomAD; ii) not intergenic; iii) within genes with known dominant disease associations in OMIM. Subsequently, we further restricted to variants with evidence of potential pathogenicity. These were variants that were either designated as Pathogenic in Clinvar, or had a CADD Phred score greater than 20, or a spliceAI (Jaganathan et al., 2019; Walker et al., 2023) score greater than 0.2. As described in Results, these filters narrowed the list of *de novo* variants down to 3 candidate variants for manual review.

## Results

### *duoNovo*: overview of approach

*duoNovo* uses haplotype blocks consisting of phased single nucleotide variants across hundreds of kilobases (Methods) in order to determine whether a candidate variant arose on the haplotype inherited from the sequenced parent – and is thus a *de novo* variant – or not. It achieves this by evaluating the sequence similarity between pairs of haplotype blocks (Methods). Each pair consists of a proband haplotype block and a parental haplotype block. If the proband haplotype block containing the variant being tested for *de novo* status exhibits perfect similarity (excluding the candidate variant) with one of the two parental haplotype blocks, then these haplotype blocks are inferred to be identical-by-descent, which in turn implies that the candidate variant is *de novo* (Figure 1A). If, on the other hand, it is the other proband haplotype (the one not containing the candidate variant) that exhibits perfect similarity with one of the two parental haplotype blocks, then *duoNovo* infers that the haplotype containing the candidate variant was inherited from the missing biological parent (Figure 1A). While in this latter case the candidate variant may still be *de novo*, this is not possible to determine from the available duo.

**Figure 1.**
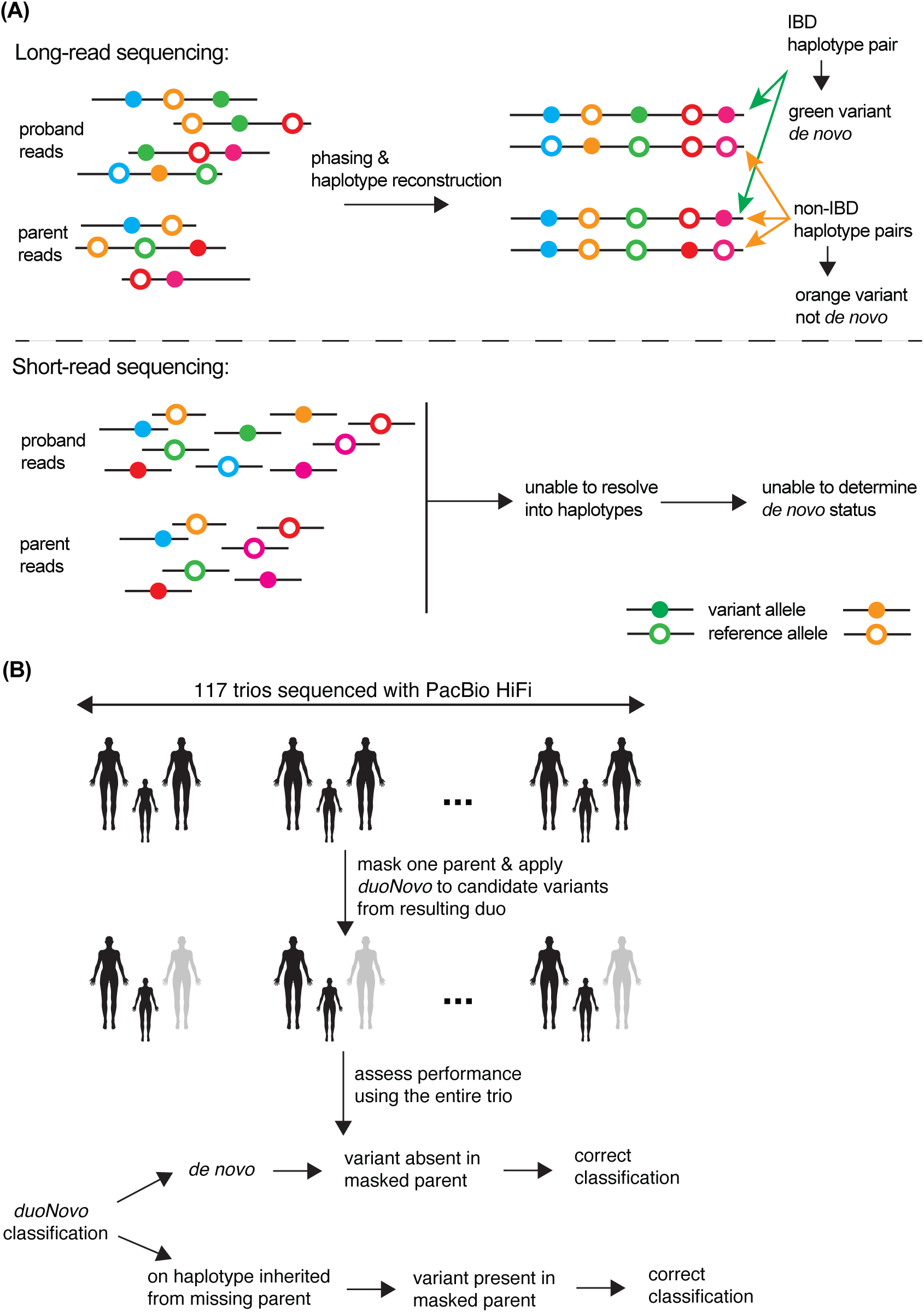
Detection of *de novo* variants from duos with *duoNovo*: conceptual basis and overview of strategy for performance testing. (A) The long reads produced by PacBio HiFi sequencing enable the read-backed phasing of variants and the subsequent reconstruction of haplotypes. *duoNovo* leverages this to identify pairs of proband-parent haplotypes that are identical-by-descent based on their sequence similarity. It then classifies candidate variants (heterozygous in the proband; absent in the parent) on such identical-by-descent haplotypes as *de novo* (orange variant in the cartoon figure; see also Methods). On the other hand, candidate variants on haplotypes not identical-by-descent with any of the two parent haplotypes (green variant in the cartoon figure) are inferred to have arisen on a haplotype inherited from the non-sequenced parent, in which case ascertaining their *de novo* status is not possible. This approach would not be feasible with short-read sequencing, as the short length of reads would preclude phasing and haplotype reconstruction. **(B)** We sequenced 117 trios with PacBio HiFi sequencing (Methods) and constructed father-proband and mother-proband duos by masking the mothers or the fathers, respectively. We then applied *duoNovo* to candidate variants from each duo, and the resulting variant classifications were evaluated based on the classifications that one would have obtained by having access to the entire trio.

### *duoNovo* has near-perfect accuracy among variants absent from gnomAD

To assess the performance of *duoNovo*, we sequenced 117 trios using PacBio HiFi LRS to an average depth of ∼ 34-fold across all individuals (SD 4.4), obtaining approximately 16.7 kb-long reads on average (SD ∼ 2kb; Methods).

We started by applying *duoNovo* to each of the 234 duos (117 father-proband duos and 117 mother-proband duos), treating either the mother or the father as the missing parent (Figure 1B). We subjected each candidate variant (heterozygous phased variant call in the proband; homozygous reference call in the parent) to strict quality control based on attributes including sequencing depth and genotype quality.

To evaluate *duoNovo*’s performance, we first sought to mimic a realistic rare disease diagnostic setting, where putative pathogenic variants have been filtered based on their frequency in control population databases. To this end, we focused on candidate variants that are absent from gnomAD v4.1 (Karczewski et al., 2020), and are thus enriched for rare pathogenic alleles. To evaluate the accuracy of each classification, we examined the missing parental sequence. For each variant that *duoNovo* classified as *de novo* from the duo, we determined that the classification is correct if the variant is absent in the missing parent (Figure 1B). We note that this a conservative assessment, as a variant can still be *de novo* even if it is present in the missing parent (for example, at recurrently mutated sites).

*duoNovo* achieved very high accuracy. Collectively, 2032 variants were classified as *de novo* from father-proband duos, among a total of 2,258,192 candidate variants. From mother-proband duos, among a total of 2,219,058 candidate variants, 531 variants were classified as *de novo*. The average (per duo) positive predictive value is 99.8% in father-proband duos, and 96.9% in mother-proband duos (Figure 2A). When applying a more stringent genotype quality threshold (GQ > 40 as opposed to > 30), we found that almost all of the false positives disappear and the positive predictive value increases to 100% in all duos except two (one father-proband and one mother-proband duo). This suggests that many of the apparent false positive *de novo* classifications are in fact genotyping errors.

**Figure 2.**
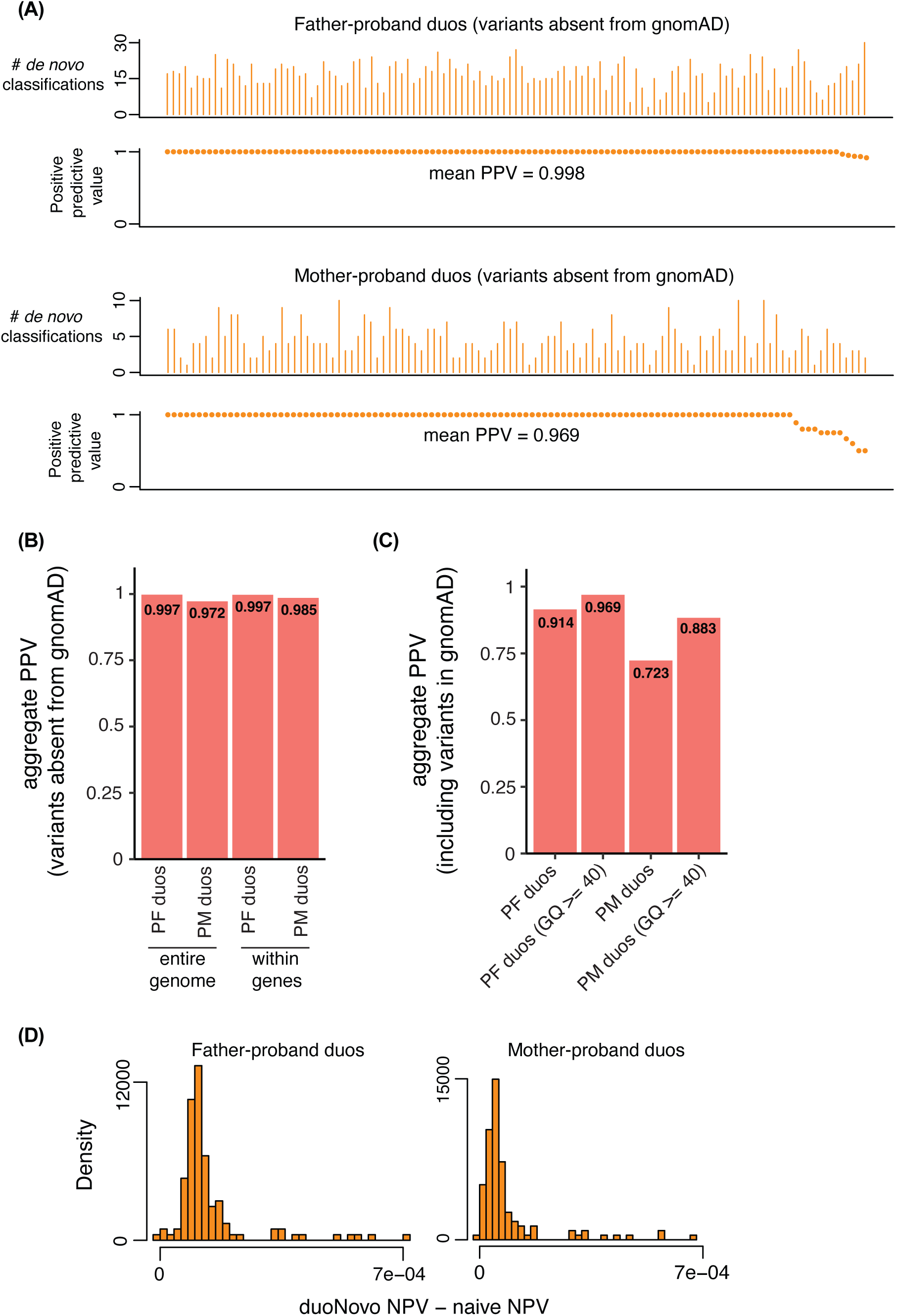
*duoNovo* has high positive and negative predictive value. **(A)** The positive predictive value (PPV) of *duoNovo* (bottom panel y axis) and the number of variants classified as *de novo* (top panel y axis). Both are calculated after first restricted to candidate variants absent from gnomAD (v4.1) Each point and its associated vertical bar above correspond to a duo. Note that duos are ordered on the x axis in decreasing PPV, and thus the order is different between the father-proband and mother-proband duos. In 5 mother-proband duos, there were no *de novo* classifications among variants absent from gnomAD, hence these duos are not visualized. **(B)** Aggregate (across all duos) PPV, calculated among all candidate variants absent from gnomAD. Shown separately when calculated using variants across the entire genome vs variants within genes only (exons and introns). **(C)** Aggregate (across all duos) PPV, calculated among all candidate variants (including these present in gnomAD). Shown separately when a more stringent genotype quality threshold (GQ ≥ 40 instead of ≥ 30) is applied in order for candidate variants to be evaluated. **(D)** Histograms showing the distribution of the differences between the negative predictive value (NPV) of *duoNovo* and the NPV of the naive baseline approach (classifying every variant as non-*de novo*). Almost all differences are positive, indicating the superior NPV of *duoNovo*.

Notably, in both father-proband and mother-proband duos, the positive predictive value remains at least as high when focusing specifically on variants within genes (exonic or intronic regions), which are the most likely to be clinically relevant (Figure 2B).

### *duoNovo*’s classifications recapitulate the known age and parent of origin effects

To gain further insight into *duoNovo*’s performance, we next examined its *de novo* classifications when testing all variants that passed QC filters (regardless of gnomAD frequency). On average, *duoNovo* classified 26 variants (range 9 - 51) as *de novo* from father-proband duos. Reassuringly, the number of *de novo* classifications positively correlates with father’s age (Supplemental Figure S1; Poisson regression *p* = 7.04 · 10^−9^). The aggregate positive predictive value of *duoNovo* when pooling classification counts across all father-proband duos is 91.4% (Figure 2C), and increases to 96.9% when restricting to candidate variants with GQ > 40 (Figure 2C; Methods).

In mother-proband duos, *duoNovo* classified an average of 9 variants as *de novo*. With the exception of one trio, *duoNovo* always detected more *de novo* variants in father-proband duos compared to mother-proband duos, with the median ratio of paternally to maternally derived *de novo* classified variants equal to 3 (Supplemental Figure S2). This is consistent with the elevated contribution of the paternal germline to the pool of *de novo* variants (Jónsson et al., 2017; Kong et al., 2012; Sasani et al., 2019), and can also explain why the positive predictive value in mother-proband duos is lower compared to father-proband duos (aggregate PPV 72.3%; increases to 88.3% when restricting to candidate variants with GQ > 40; Figure 2C).

In both father-proband and mother-proband duos, we observed no major difference in the positive predictive value when the proband was of European vs non-European ancestry (with slightly higher PPV in duos with non-European ancestry probands; Supplemental Figure S3; Methods).

### Variants classified as *de novo* fall into expected mutation subtypes

In agreement with previously reported mutational patterns among *de novo* variants (Kessler et al., 2020; Shojaeisaadi et al., 2024), we found that the single nucleotide variants *duoNovo* classified as *de novo* are mostly C>T and T>C substitutions (Supplemental Figure S4), with the C>T substitutions occurring both within and outside the CpG context. There was no statistically significant difference in the proportion of the 6 mutation types among the *de novo* classifications from father-proband duos (2906 single nucleotide variants), and those from mother-proband duos (1009 single nucleotide variants) (Bonferroni adjusted *p >* 0.05 for all types; Supplemental Figure S4).

### *duoNovo* has very high negative predictive value

The negative predictive value of any sensible approach to classifying variants as *de novo* is expected to be high because *de novo* variants are extremely rare. To provide a fair assessment, we chose to compare *duoNovo* to the “naive baseline” approach: classifying every variant as non-*de novo*. We found that *duoNovo* has a higher negative predictive value in all duos except one father-proband and one mother-proband duo (Figure 2D; Supplemental Figure S5).

### *duoNovo* very rarely classifies reference alleles as *de novo*

Our results so far focus on candidate alternative alleles (1|0 or 0|1 proband genotype; 0/0 parental genotype). As an additional check of *duoNovo*’s performance, we compared the percentage of candidate alternative alleles classified as *de novo* to the corresponding percentage for candidate reference alleles (1|0 or 0|1 proband genotype; 1/1 parental genotype). Collectively, alternative candidate alleles were ∼ 35 times more likely to be classified as *de novo* compared to reference candidate alleles in father-proband duos, and ∼ 12 times in mother-proband duos. In 78 out of the 117 father-proband duos and 75 out of the 117 mother-proband duos, no reference alleles were classified as *de novo* (Supplemental Figure S6). This further supports the accuracy of our *de novo* classifications, since the reference allele is common in the population and thus candidate reference alleles have a very low prior probability of being *de novo*.

### *duoNovo* has very low false positive rate

To directly quantify the genome-scale false positive rate of *duoNovo*, we subsequently restricted our attention to candidate variants which we inferred – based on the full trio information – to have been transmitted from the non-sequenced parent (reliable heterozygous phased variant call in the proband and reliable heterozygous or homozygous variant call in the non-sequenced parent; reliable homozygous reference call in the sequenced parent). We examined the fraction of these variants that are classified as *de novo* (false positive rate; equivalent to the specificity), and found that *duoNovo* has an average false positive rate across father-proband and mother-proband duos equal to 5.1e-6, and 5.8e-6, respectively (Figure 3A, B). The majority (more than 60%) of these variants were correctly classified as present on the haplotype inherited from the missing parent, with almost 40% labeled as uncertain (Figure 3A). The maximum false positive rate across all duos was 1.9e-5. As noted earlier, this is a conservative assessment since some of these variants may in fact be *de novo* even though they are present in the non-sequenced parent.

**Figure 3.**
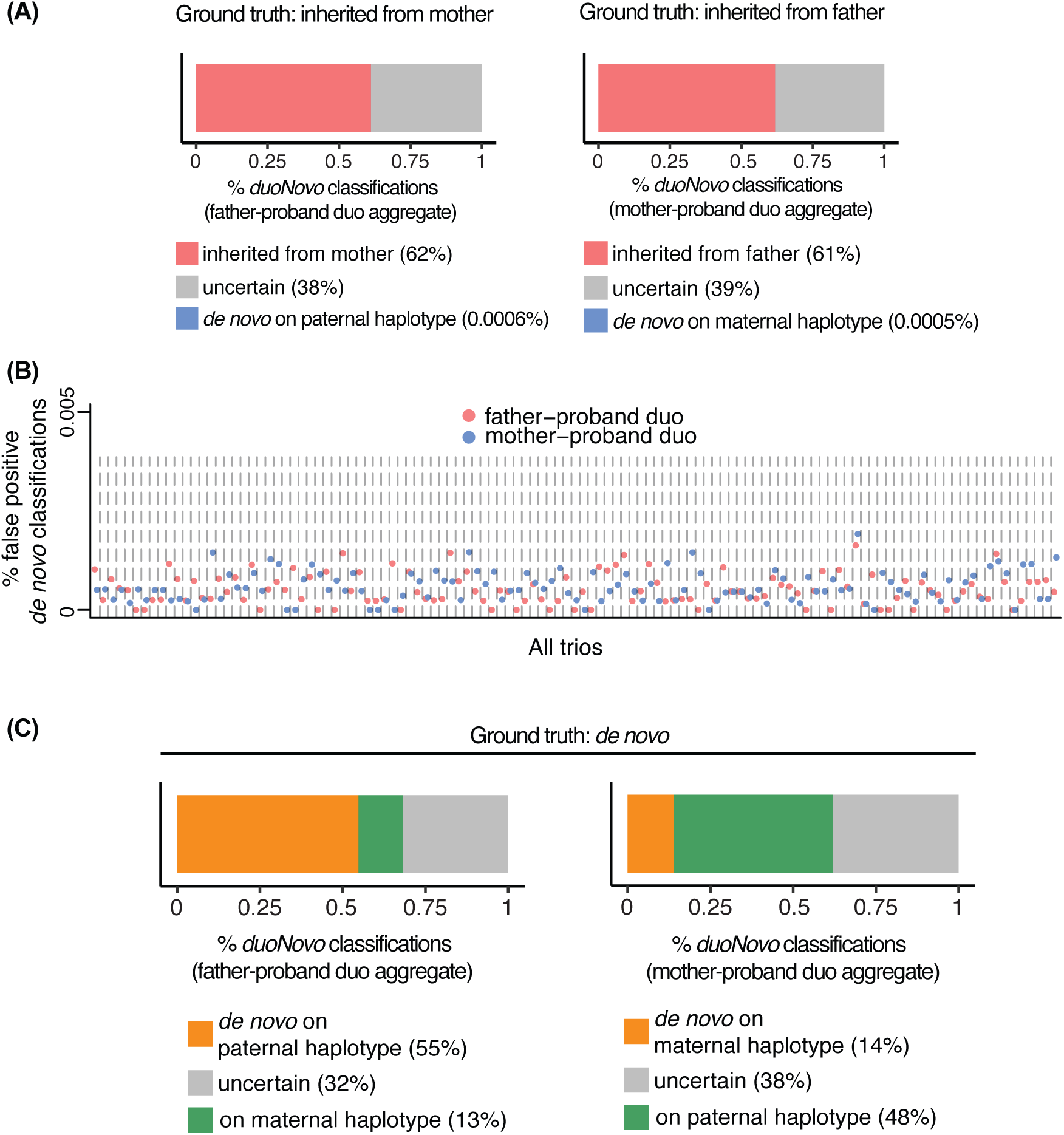
*duoNovo* has very low error rate. **(A)** The percentage of candidate variants present in the non-sequenced parent (and thus inferred to be non-*de novo*) that are classified by *duoNovo* as present on the haplotype inherited from the masked parent, *de novo* or uncertain. Percentages (x axis) were computed after pooling counts across all father-proband or mother-proband duos. Variants that did not pass QC filters required to proceed with classification (Methods), or variants within a haplotype containing multiple *de novo* classifications (which are usually artifacts; Methods) are excluded. **(B)** The false positive rate (fraction of non-*de novo* variants misclassified as *de novo*; y axis) of *duoNovo*, calculated separately for each father-proband and mother-proband duo. **(C)** The percentage of *de novo* variants detected from the entire trio (Methods) that were labeled as *de novo*, present on the haplotype inherited from the non-sequenced parent, or uncertain. Percentages (x axis) were calculated after pooling counts across all father-proband or mother-proband duos. Variants that failed QC are excluded from the percentage calculation.

As an orthogonal assessment of the false positive rate, we applied *duoNovo* to 40 swapped duos consisting of the father and the mother from the corresponding trio (that is, the proband was swapped with the parent previously treated as the non-sequenced one). From each swapped duo, we calculated the percentage of candidate variants that were classified as “*de novo*”, or as “present on the non-sequenced parent haplotype”. Either classification requires the detection of a haplotype block shared identical-by-descent between the two members of the duo. We would thus expect the percentage of classified variants from these swapped duos to be very low, since fathers and mothers share a much smaller percentage of their genome identical-by-descent compared to father-proband or mother-proband pairs. Indeed, we found that the median percentage across swapped duos is 0.03%, compared to 65% across true parent-proband duos (Supplemental Figure S7). One swapped duo stands out as an outlier, with 10.6% of variants receiving a classification (Supplemental Figure S7); this is substantially higher than all the rest, though still well below the percentage from true parent-proband duos (Supplemental Figure S7). Upon closer examination, we found that the father and mother from that swapped duo had the highest relatedness score among all father/mother pairs (Methods; Supplemental Figure S7), which could explain why *duoNovo* detected a larger fraction of identical-by-descent haplotype blocks.

### *duoNovo*’s sensitivity reflects the parent of origin effect

We next sought to characterize the sensitivity of *duoNovo*, based on a set of *de novo* variants detected using an established approach from the entire trio (hereafter referred to as trio-*de novo* variants; Methods), which we treated as the ground truth. Consistent with the aforementioned fact that most *de novo* variants are contributed by the fathers, we found that *duoNovo* collectively classified ∼ 55% of trio-*de novo* variants as *de novo* from the father-proband duos, and only ∼ 13% as *de novo* from the mother-proband duos (Figure 3C). Conversely, approximately 13% were classified as present on the maternally inherited haplotype from the father-proband duos, whereas about 48% were classified as present on the paternally inherited haplotype from the mother-proband duos (Figure 3C). 32% and 48% of variants were labeled as uncertain from the father-proband and mother-proband duos, respectively. Looking at the total fraction of trio-*de novo* variants that are classified as *de novo* (either from the father-proband or from the mother-proband duo; sensitivity), we found that the average sensitivity across all trios is 51% (Supplemental Figure S8). The remaining were all variants that were either labeled as uncertain or did not pass our QC filters; none of the trio-*de novo* variants were misclassified as non-*de novo*.

### Using siblings as surrogates for missing fathers boosts *duoNovo*’s sensitivity

The aforementioned fact that most *de novo* variants are transmitted from the paternal germline raises a practical issue pertinent to the clinical application of *duoNovo*, since most single-parent families are single-mother families. To mitigate this, we reasoned that siblings – when available – can increase *duoNovo*’s sensitivity by serving as surrogates for the missing father, since they share 50% of their genome with the father. To test this, we focused on 29 mother-proband duos where siblings were available. We first obtained the variants that *duoNovo* classified from the mother-proband duos as either present on the haplotype inherited from the missing father, or uncertain. We then tested these variants for *de novo* status from the proband-sibling-mother trio (Methods). We discovered that this leads to a 86% increase in sensitivity, from 12.8% to 23.9% (Figure 4A). This shows that the inclusion of siblings in the analysis can indeed partially compensate for the missing biological father. By contrast, and consistent with expectation, applying the same approach to proband-sibling-father trios only led to a negligible increase in the sensitivity compared to father-proband duos (57.3% compared to 54.6% without siblings; Figure 4A).

**Figure 4.**
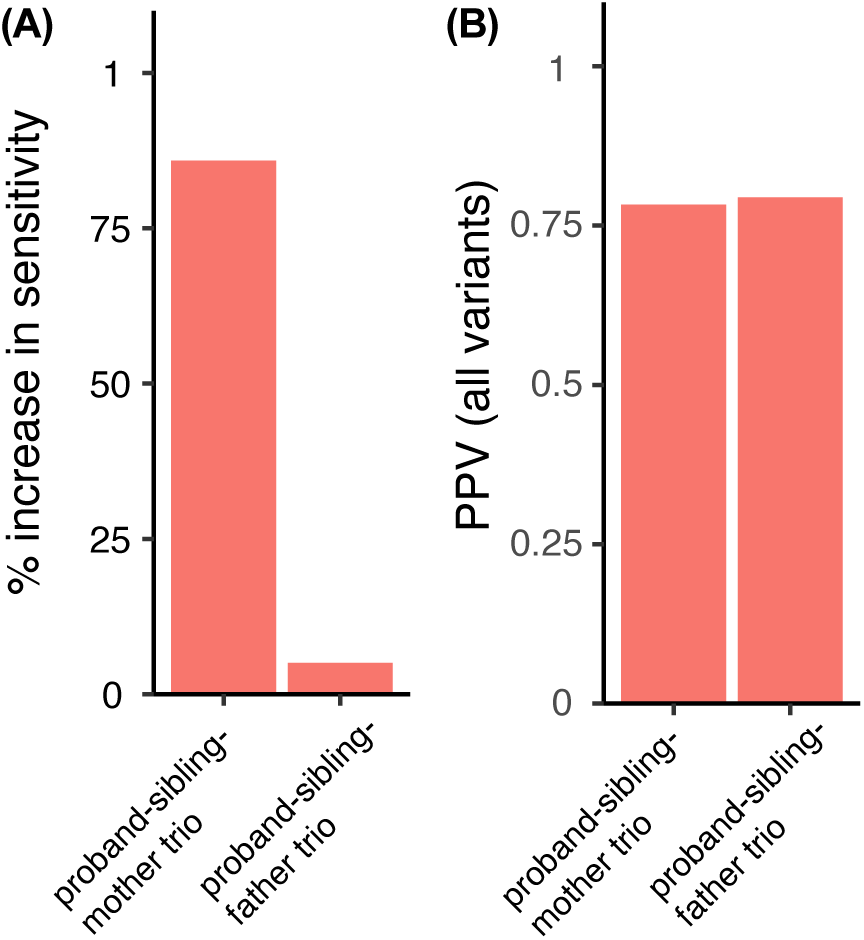
Using siblings as surrogates for the missing father increases the sensitivity of *duoNovo* from proband-mother duos. **(A)** The % increase in sensitivity when using the proband-sibling-mother or proband-sibling-father trio to classify candidate variants initially classified as either present on the haplotype inherited from the missing parent, or uncertain, from the proband-mother or proband-father duo, respectively. **(B)** The positive predictive value when classifying candidate variants using either the proband-sibling-mother or the proband-sibling-father trio. The PPV is here calculated among candidate variants obtained from the entire genome (that is, without restricting to variants initially classified as either present on the haplotype inherited from the missing parent, or uncertain, from the duos). Both **(A)** and **(B)** depict aggregate percentages, calculated after pooling classification counts across all 29 mother-proband and father-proband duos with siblings available (see Methods for details).

We also examined the genome-scale positive predictive value of this approach. We obtained candidate variants based on genotype (heterozygous variant call in the proband; homozygous reference call in both the sibling and the parent), regardless of how these variants were classified from the proband-mother or proband-father duos. We classified these candidate variants directly using the corresponding proband-sibling-mother or proband-sibling-father trio, and then examined the missing parent’s genotype at the positions of *de novo* classifications to determine their correctness. We found that the PPV is equal to 78.2% and 79.4% from proband-sibling-mother and proband-sibling-father trios, respectively (Figure 4B).

### *duoNovo* makes no erroneous classifications among a set of manually curated, clinically relevant variants

We next assessed *duoNovo*’s performance on a set of variants with known inheritance status that were manually curated as putatively clinically relevant in the UCI-GREGoR case set, a cohort of probands suspected to have a Mendelian disorder but who remained undiagnosed despite prior genetic testing.

We first focused on 22 *de novo* variants, identified from 22 trios. We applied *duoNovo* to both the father-proband and the mother-proband duo corresponding to each trio, with settings adjusted for targeted variant analysis – which account for the higher prior probability of candidate variants deemed to be of clinical interest after manual curation as opposed to the lower prior of candidate variants identified genome-wide solely based on genotype (Methods). We classified 12 of the 22 variants as *de novo*, equivalent to a sensitivity of ∼ 55% (Table 1). Unsurprisingly, 11 of the 12 *de novo* classifications came from the corresponding father-proband duo. Of the remaining variants, 5 were classified as on the missing parent haplotype from one duo but were either uncertain or failed QC (before proceeding to classification) from the other duo, 4 were classified as uncertain from both duos, and 1 was classified as uncertain from one duo but failed QC from the other duo. The predominant reason that variants failed QC in this analysis was that they were not resolved into a haplotype block for *duoNovo* to be able to classify them. Importantly, *duoNovo* did not misclassify any of these variants.

**Table 1.**
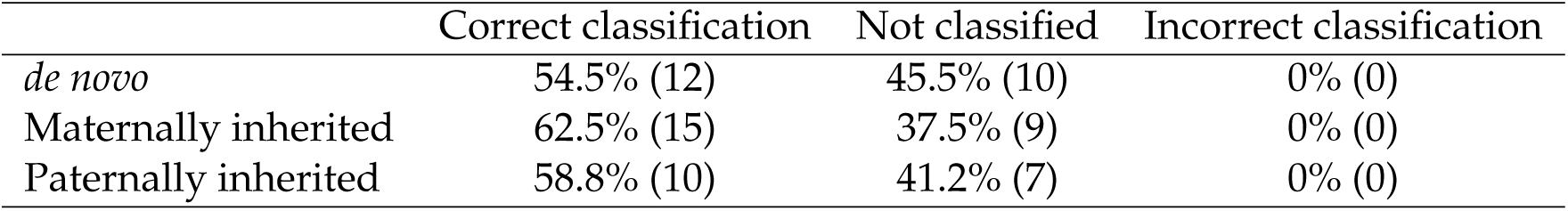
Performance of *duoNovo* on manually curated variants of clinical relevance from the UCI-GREGoR case set. The *de novo* variants were tested from both duos. The maternally inherited variants were tested from the father-proband duo, while the paternally inherited variants were tested from the mother-proband duo.

|  | Correct classification | Not classified | Incorrect classification |
| --- | --- | --- | --- |
| *de novo* | 54.5% (12) | 45.5% (10) | 0% (0) |
| Maternally inherited | 62.5% (15) | 37.5% (9) | 0% (0) |
| Paternally inherited | 58.8% (10) | 41.2% (7) | 0% (0) |

We also examined 41 non-*de novo* variants for which we knew the parent of origin (17 paternal and 24 maternal). When applied to the duo consisting of the proband and the non-transmitting parent, *duoNovo* correctly classified 25 out of the 41 variants (61%) as present on the other parent’s haplotype (Table 1). In all the remaining cases, the variant was either labeled as uncertain or failed QC; none of these variants were misclassified as *de novo*.

### *duoNovo* prioritizes a *de novo* variant in a cohort of undiagnosed rare disease cases

Finally, we applied *duoNovo* to 74 duos (52 mother-proband; 22 father-proband) from the UCI-GREGoR case set; several of these probands still remained undiagnosed even after enrollment in the GREGoR study. Across all duos, *duoNovo* classified 917 variants as *de novo*. After additional filtering to narrow this list down to variants likely to be clinically relevant (Methods), we identified 3 candidate variants, two of which were previously reported as Pathogenic in ClinVar. The first is an insertion in *RNU4-2* (n.64 65insT) that was identified in a mother-proband duo. This variant explains the proband’s severe developmental delay and microcephaly, and the maternal origin of the variant is consistent with the recent discovery that it is recurrent *de novo* variant on the maternally transmitted haplotype in cases of neurodevelopmental delay (Chen et al., 2024; Greene et al., 2024). The other *de novo* pathogenic variant – identified in a father-proband duo – is a variant in *ACTA1* (NM 001100.4:exon3:c.C282A:p.N94K) which explains the proband’s congenital myopathy (MIM: 102610). In addition to these two pathogenic variants, we discovered a *de novo* intronic variant in *PHIP* (c.4206+3A>G; Supplemental Figure S9) in a proband with developmental delays and syndromic features. This variant, identified from a mother-proband duo, has a spliceAI score of 0.27, suggesting an impact on splicing (Walker et al., 2023). Haploinsufficiency of *PHIP* causes Chung-Jansen Syndrome (MIM: 617991), a disorder associated with developmental delays, intellectual disability, and distinct facial features (Jansen et al., 2018; Webster et al., 2016). While this variant is still labeled as VUS because of the nature of the phenotype and further functional studies are needed to confirm its pathogenicity, *duoNovo* led to the prioritization of this variant by uncovering its previously unknown *de novo* status.

We also specifically looked at 10 variants that were already curated as putatively clinically relevant, but were identified from duos and as such had uncertain *de novo* status. For these variants, we used the targeted *duoNovo* settings (Methods). Of those variants, 8 were identified from mother-proband duos and 2 were identified from father-proband duos. After applying *duoNovo*, 7 of these variants were classified as present on the missing parent’s haplotype (6 from mother-proband duos; 1 from a father-proband duo), 2 were uncertain, and one failed QC.

## Discussion

We have developed and extensively evaluated a method that enables the identification of *de novo* variants from parent-proband duos. Our approach is simple, and leverages the unique ability of LRS to both accurately detect and phase variants across large genomic segments.

Although LRS is currently more expensive than SRS, the cost differential between LRS and SRS has significantly decreased over the past decade (McCombie and McPherson, 2019), and that trend is likely to continue as a result of continued improvements in technology. It is thus imperative to characterize the diagnostic benefit of LRS over SRS, as accurately and comprehensively as possible. While an extensive body of work has established that LRS can detect variation that is undetectable with SRS (such as complex structural variants; Cohen et al. (2022), Dominguez Gonzalez et al. (2025), Mastrorosa et al. (2023), Merker et al. (2018), Miller et al. (2021), Mizuguchi et al. (2021), Negi et al. (2025), and Xie et al. (2020)), *duoNovo* illustrates that LRS can also enable the interpretation of variants that can be detected with SRS, but are hard-to-interpret because of unknown *de novo* status. Importantly, the ability to ascertain *de novo* status from single-parent genetic testing promises to address an important source of inequity in result interpretation and diagnostic outcomes.

Conceptually, *duoNovo* is based on the recognition that phasing can provide information about haplotype transmission, and thus reveal variant inheritance, without access to both biological parent genotypes (see also Steyaert et al. (2024)). As such, it has the potential to improve variant interpretation in cases where sequencing both parents is not feasible, addressing a critical unmet need in clinical genetic testing. An inherent limitation of our approach is that it can only detect *de novo* variants derived from the germline of the available sequenced parent. Given that the paternal germline contributes approximately 3-4 times as many *de novo* single nucleotide variants than the maternal germline, whereas most single-parent families are single-mother families (Jónsson et al., 2017; Kong et al., 2012; Sasani et al., 2019), this will limit the yield in assigning *de novo* status to variants identified clinically. We show that this challenge can be partially addressed, however, if siblings are available and are used as surrogates for the missing father. A potential source of false positives that one must be aware of when using the sibling trio-based approach are near-homozygous haplotypes in the missing parent; haplotypes differing only in the presence of the candidate variant. If the copy of the haplotype without the variant has been transmitted to the sibling, then the variant will be misclassified as *de novo* from the sibling-based trio approach.

In this study, we have focused on single nucleotide variants and indels. However, we anticipate that *duoNovo* is going to be similarly useful for additional variant types which LRS is capable of reliably detecting, such as structural variants and short tandem repeats (Beyter et al., 2021; Chaisson et al., 2015; Figueroa et al., 2024; Logsdon et al., 2020; Sedlazeck et al., 2018; Smolka et al., 2024). Furthermore, although we used PacBio HiFi LRS to develop and test our method, *duoNovo* is generally applicable to any sequencing platform that generates accurate variant calls and enables read-backed haplotype reconstruction. Finally, we note that our current Methods of *duoNovo* uses certain hard thresholds for parameters relevant to variant call reliability such as sequencing depth and genotype quality. These thresholds are supported by our extensive sensitivity analysis, though as mentioned we consider it sensible to use more lenient thresholds for targeted variant analyses as opposed to genome-scale analyses, since the former focuses on variants that have already undergone manual curation and thus have a higher-than-baseline prior probability of being *de novo*. An alternative approach to the one we have taken here is to treat the thresholds as learnable parameters in a machine learning model trained to predict the *de novo* status of a variant. We view this as a very promising avenue for a future extension of *duoNovo*, though care must be taken when choosing the labeled examples to train the model on, and to deal with issues related to class imbalance (*de novo* variants being very rare).

In summary, *duoNovo* is the first systematic method that can identify *de novo* variants from duos with high accuracy at the genome scale. It has the potential to transform the diagnostic yield of genetic testing for millions of single-parent families, and represents an example where long-read sequencing can provide clear benefit compared to short-read sequencing in the clinical diagnostic setting. We anticipate that its Methods and free availability as an R package will facilitate its use and adoption by the community.

## Ethics approval and consent to participate

All individuals consented to provide samples for genetic testing to the Pediatrics Mendelian Genomics Research Center (part of the GREGoR consortium). The study was approved by Children’s National Hospital IRB (IRB #PRO00015852).

## Availability of data and materials

Sequencing data are available on AnVIL as part of the GREGoR Consortium data release (https://gregorconsortium.org/data). *duoNovo* is freely available for installation on https://github.com/sbergercnmc/duonovo.

Operating system(s): Platform independent

Programming language: R and Bash Script

Other requirements: VariantAnnotation R Package

License: MIT

Any restrictions to use by non-academics: none

## Author contributions

SIB conceived of the study. LB and SIB designed and implemented *duoNovo*. LB and SIB processed and analyzed data. LB visualized data. CL, CF, PB, SB, BL, JH, KB, RK and WR performed the long-read sequencing and initial post-sequencing processing. LB, GP and SIB organized participant information. LB wrote the paper, which was then revised and edited by LB and SIB, with critical input from ED, GP, WR, JL, HAK, CX, EV. All authors reviewed the data and approved of the final manuscript.

## Data Availability

Sequencing data are available on AnVIL as part of the GREGoR Consortium data release.

https://gregorconsortium.org/data

## List of abbreviations

LRS: Long-read sequencing
SRS: Short-read sequencing
PPV: Postivive Predictive Value
QC: Quality Control

## Supplemental Figures

**Supplemental Figure S1.**
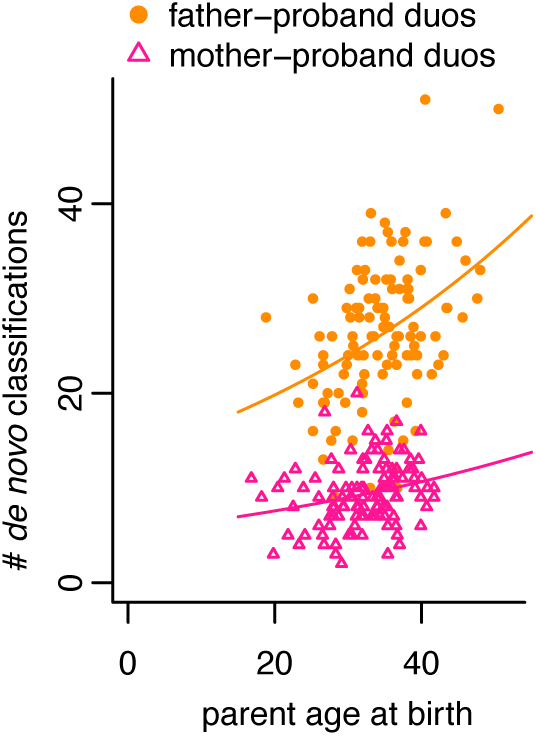
Paternal age positively correlates with the number of *de novo* classifications from father-proband duos. Each point corresponds to a duo. Trend lines were plotted after fitting Poisson regression models using the log link. As seen in previous studies (Sasani et al., 2019), there is also a significant, but weaker, effect of maternal age.

**Supplemental Figure S2.**
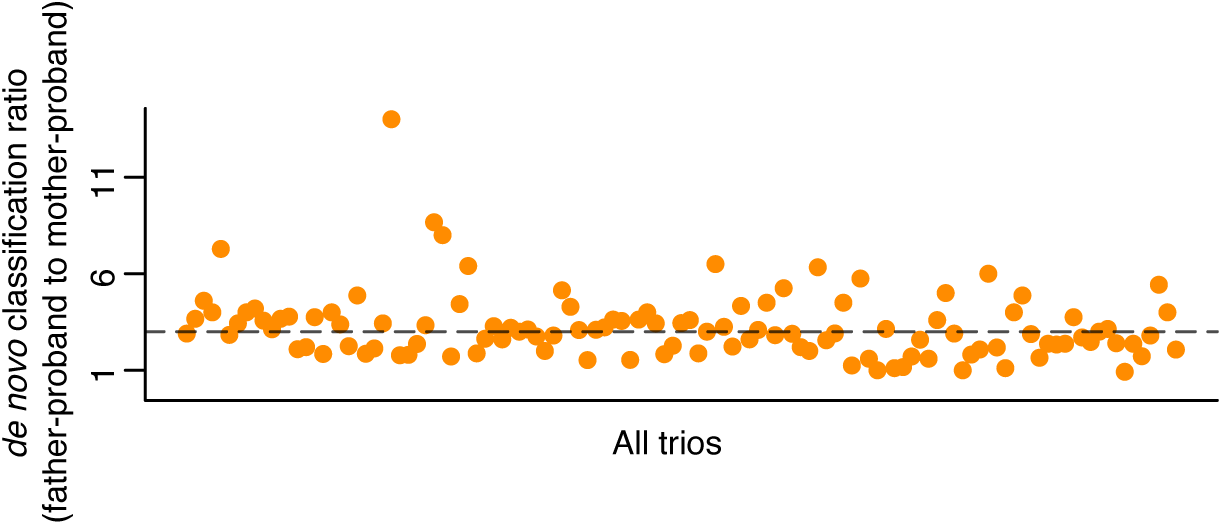
*duoNovo* detects more *de novo* variants from father-proband duos compared to mother-proband duos. Each point corresponds to a trio and its position on the y axis corresponds to the ratio of the number of *de novo* classifications from the father-proband duo to the number of *de novo* classifications from the mother-proband duo. The dashed horizontal line corresponds to the median across all trios.

**Supplemental Figure S3.**
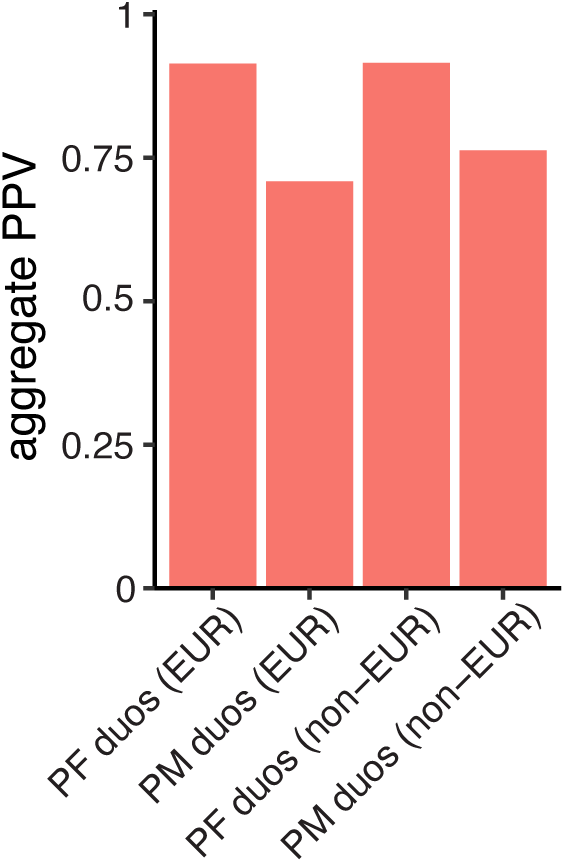
No major difference in the positive predictive value of *duoNovo* in probands of European vs non-European ancestry. Aggregate (across all duos) positive predictive value, shown separately for duos with probands of European versus non-European ancestry. Ancestry was calculated using Somalier (Pedersen et al., 2020). Non-European ancestry groups include African (2 probands), Admixed American (20 probands), East Asian (3 probands), and South Asian (5 probands).

**Supplemental Figure S4.**
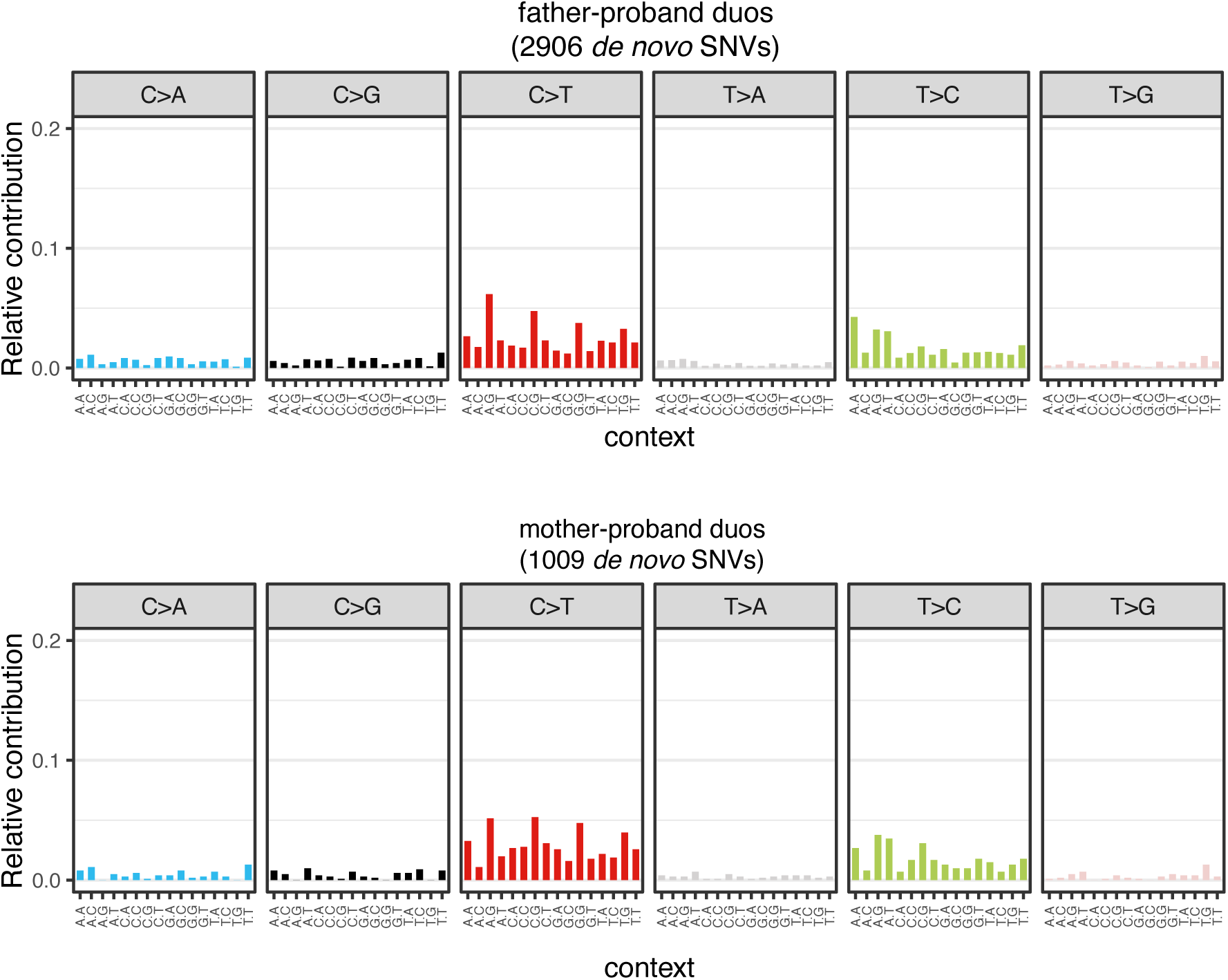
The de novo single nucleotide variants identified by duoNovo fall into expected mutation subtypes. The relative contributions were calculated after pooling all single nucleotide variants classified as de novo from the father-proband duos (top) and the mother-proband duos (bottom).

**Supplemental Figure S5.**
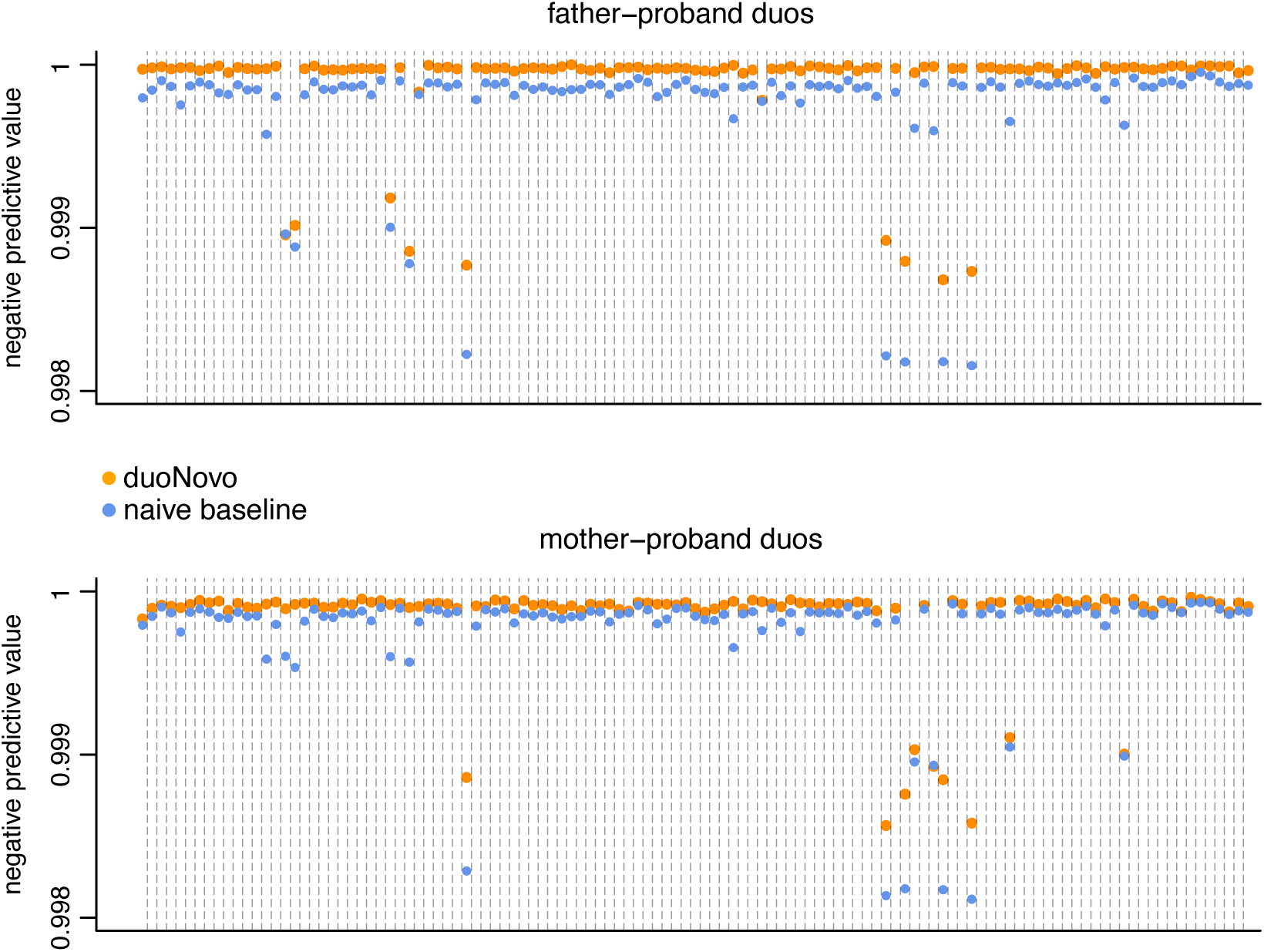
*duoNovo* has higher negative predictive value than the naive baseline (classifying every variant as non-*de novo*). The negative predictive value (y axis) of *duoNovo* (orange points) and of the naive baseline approach (blue points) for each father-proband and mother-proband duo.

**Supplemental Figure S6.**
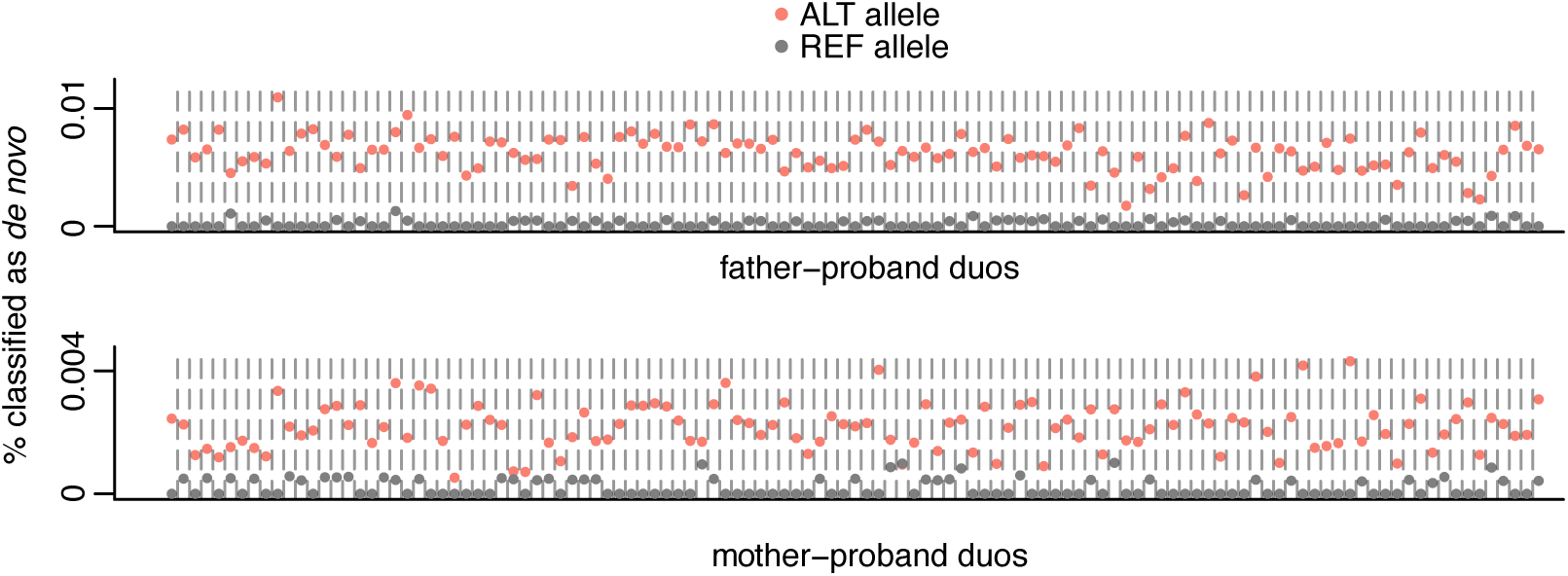
*duoNovo* very rarely classifies candidate reference alleles as *de novo*. Candidate alternative (ALT) alleles were identified by finding positions where the proband had the “1|0” or “0|1” genotype and the parent had the “0/0” genotype. Conversely, candidate reference (REF) alleles were identified by finding positions where the proband had the “1|0” or “0|1” genotype and the parent had the “1/1” genotype. Positions were either the proband or the parent failed QC filters (Methods) were excluded.

**Supplemental Figure S7.**
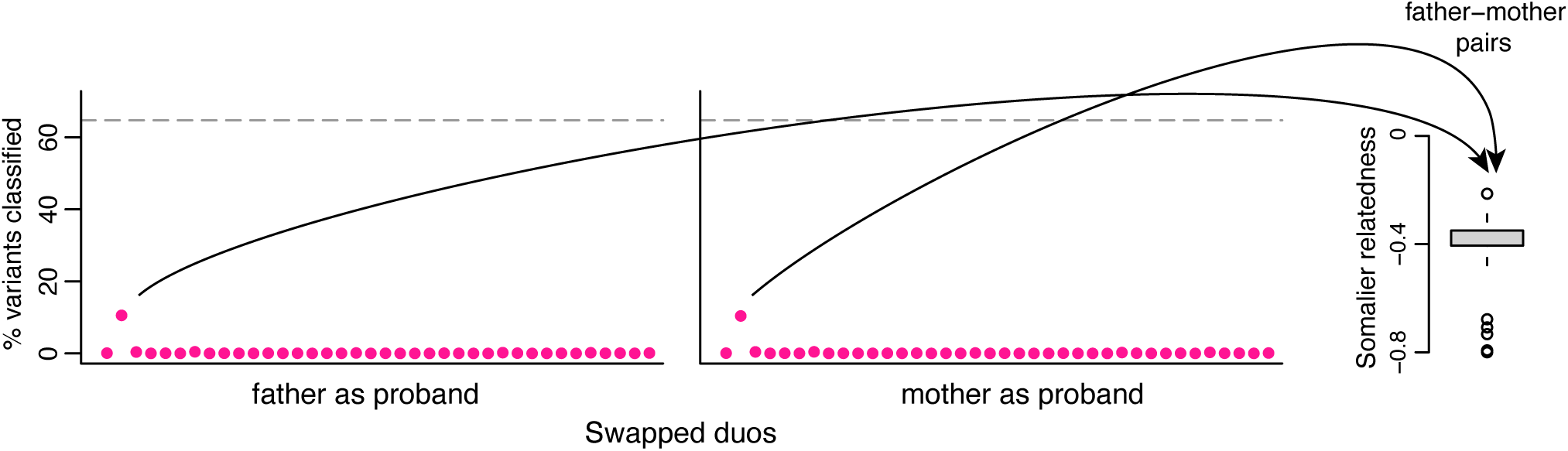
*duoNovo* does not classify variants from swapped duos. Each point corresponds to a swapped duo, and its position on the y axis indicates the percentage of variants that received a classification (either *de novo* or on the non-sequenced parent haplotype). Each swapped duo consists of the two parents instead of a parent and the proband; *duoNovo* was applied to each swapped duo by either treating the father as the proband (left) or the mother as the proband (right). The dashed horizontal line corresponds to the median percentage of variants that received a classification across all regular (father-proband or mother-proband) duos. The rightmost panel depicts the distribution of relatedness coefficients between all the fathers and mothers (each point corresponds to a father-mother pair), showing that the outlier swapped duo is the one with the highest relatedness coefficient.

**Supplemental Figure S8.**
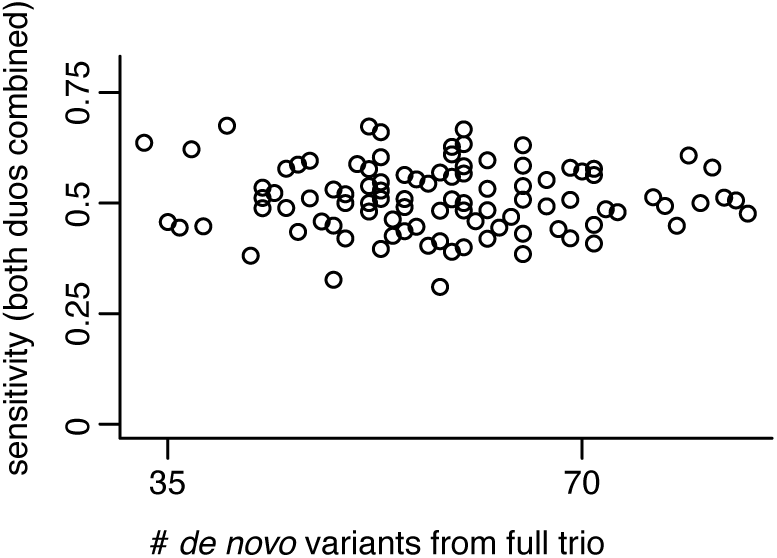
The relationship between *duoNovo*’s sensitivity (when combining *de novo* variants detected from father-proband and mother-proband duos) and the number of ground truth *de novo* variants detected from the full trio. Each point corresponds to a trio.

**Supplemental Figure S9.**
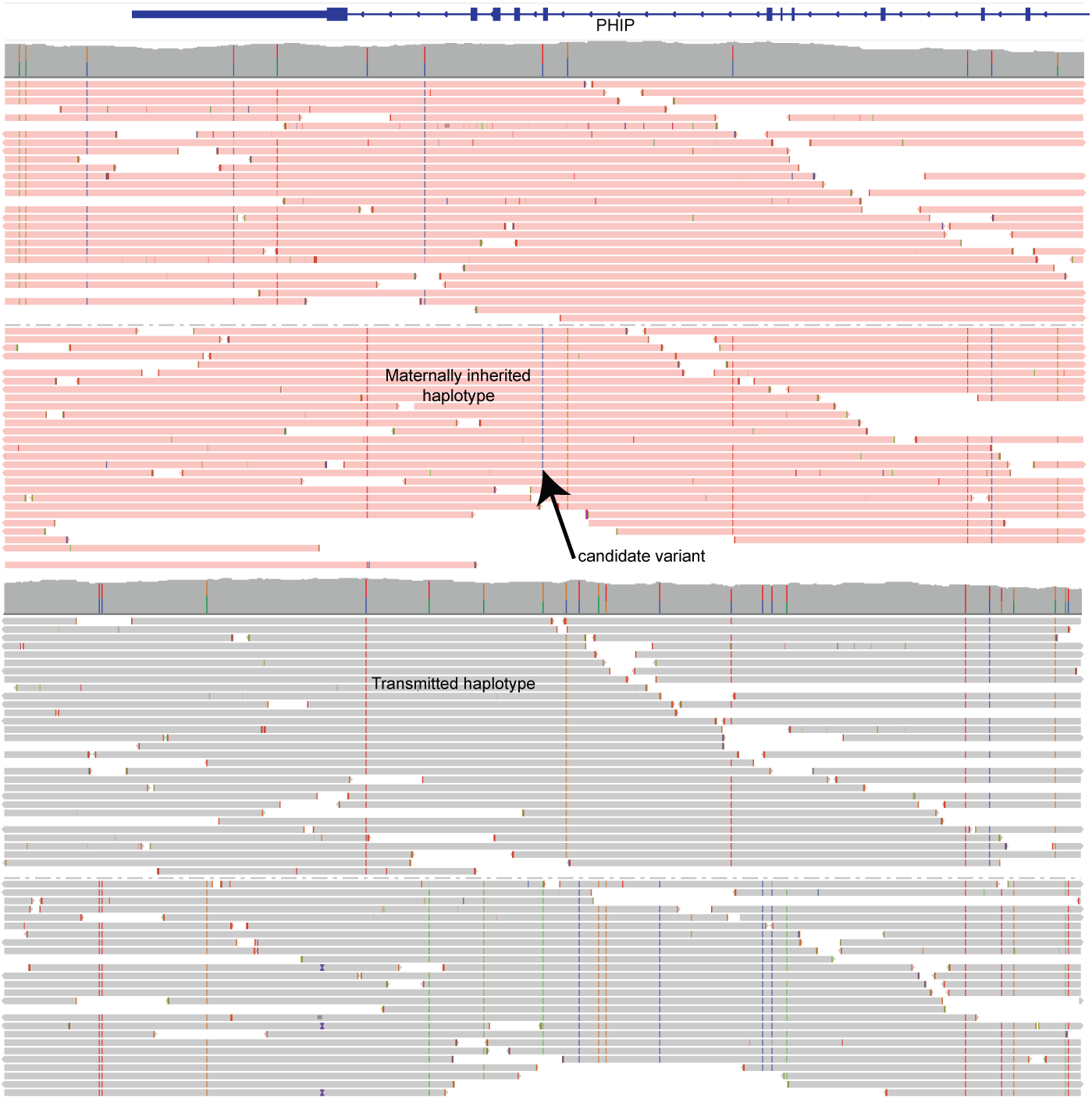
IGV snapshot of the sequencing reads from the proband harboring the *PHIP* intronic variant classified as *de novo* and the mother. Proband reads are shown in pink at the top and are grouped according to phase; the dashed line in the middle separates the two groups. Similarly, the maternal sequencing reads are shown in gray at the bottom grouped according to phase, with the dashed gray line separating the two groups.

**Supplemental Figure S10.**
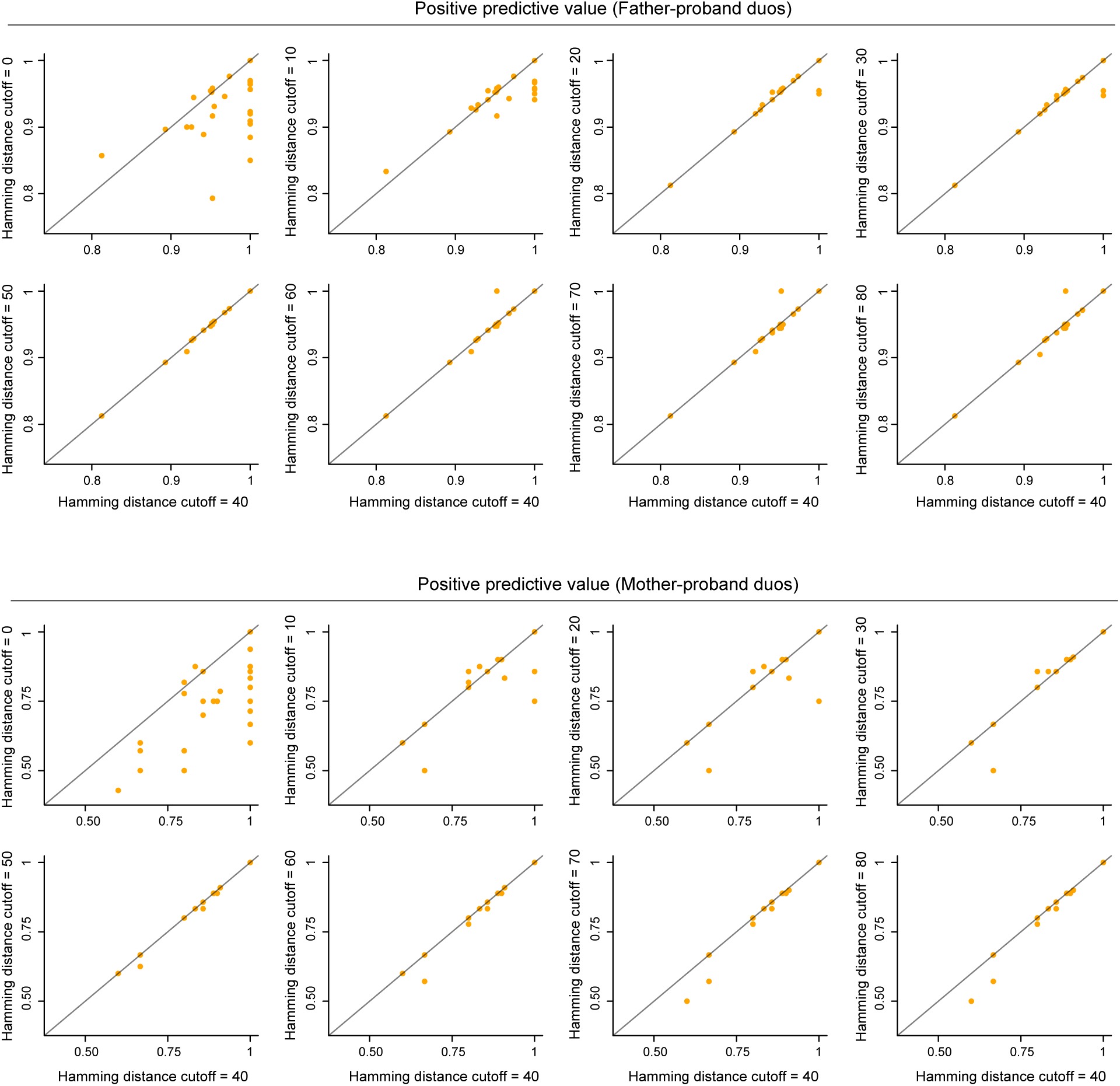
The impact of the Hamming distance threshold for determining dissimilarity between a pair of haplotype blocks on the positive predictive value. Each point corresponds to a duo. The x-axis shows the positive predictive value (PPV) when using a Hamming distance threshold of 40 (default), and the y axis shows the PPV when using different Hamming distance thresholds. There is some degree of overplotting, due to duos that have the same or highly similar PPV.

**Supplemental Figure S11.**
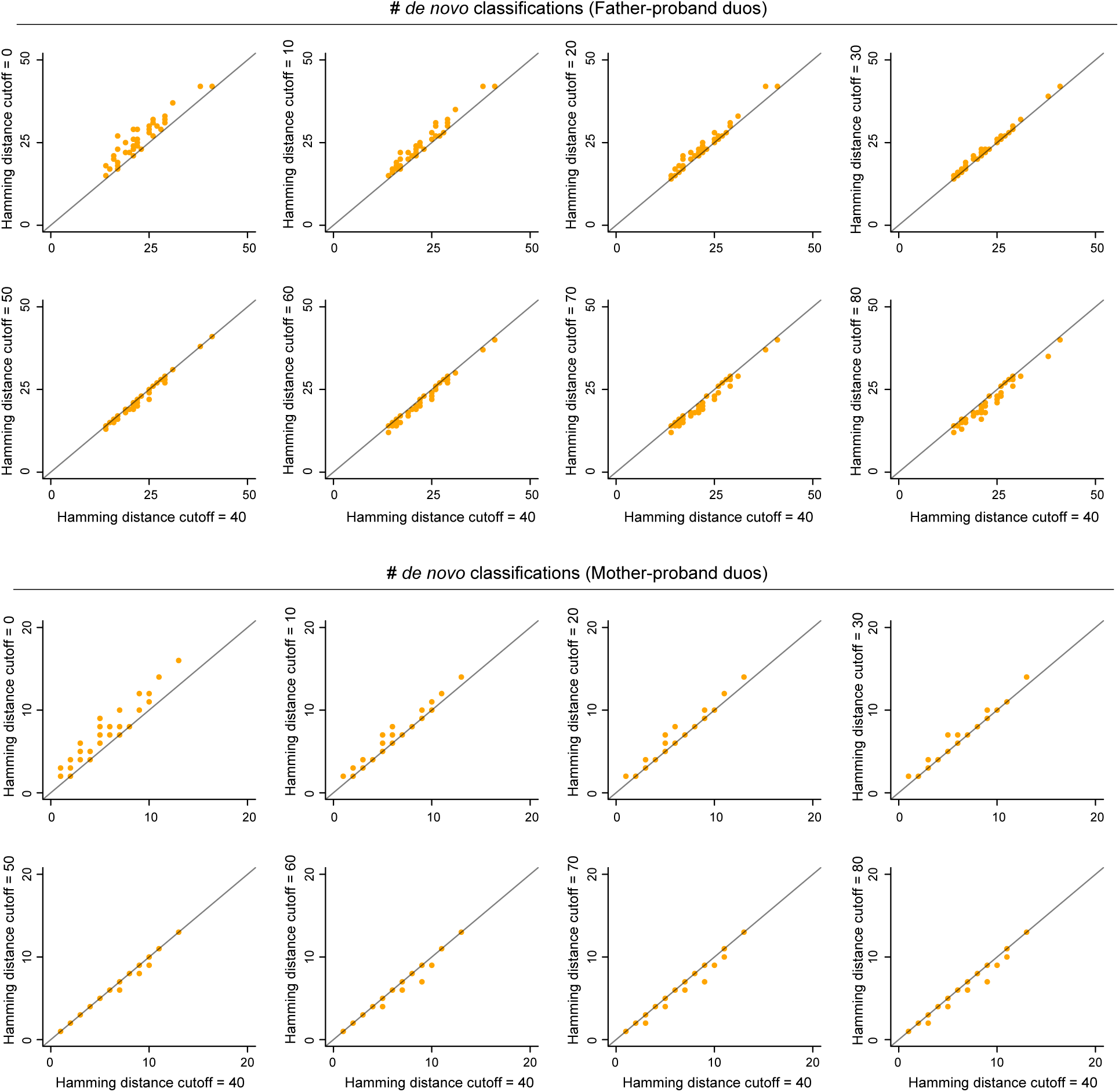
The impact of the Hamming distance threshold for determining dissimilarity between a pair of haplotype blocks on the number of *de novo* classifications. Like Supplemental Figure S10, but depicting the number of *de novo* classifications instead of the PPV.

**Supplemental Figure S12.**
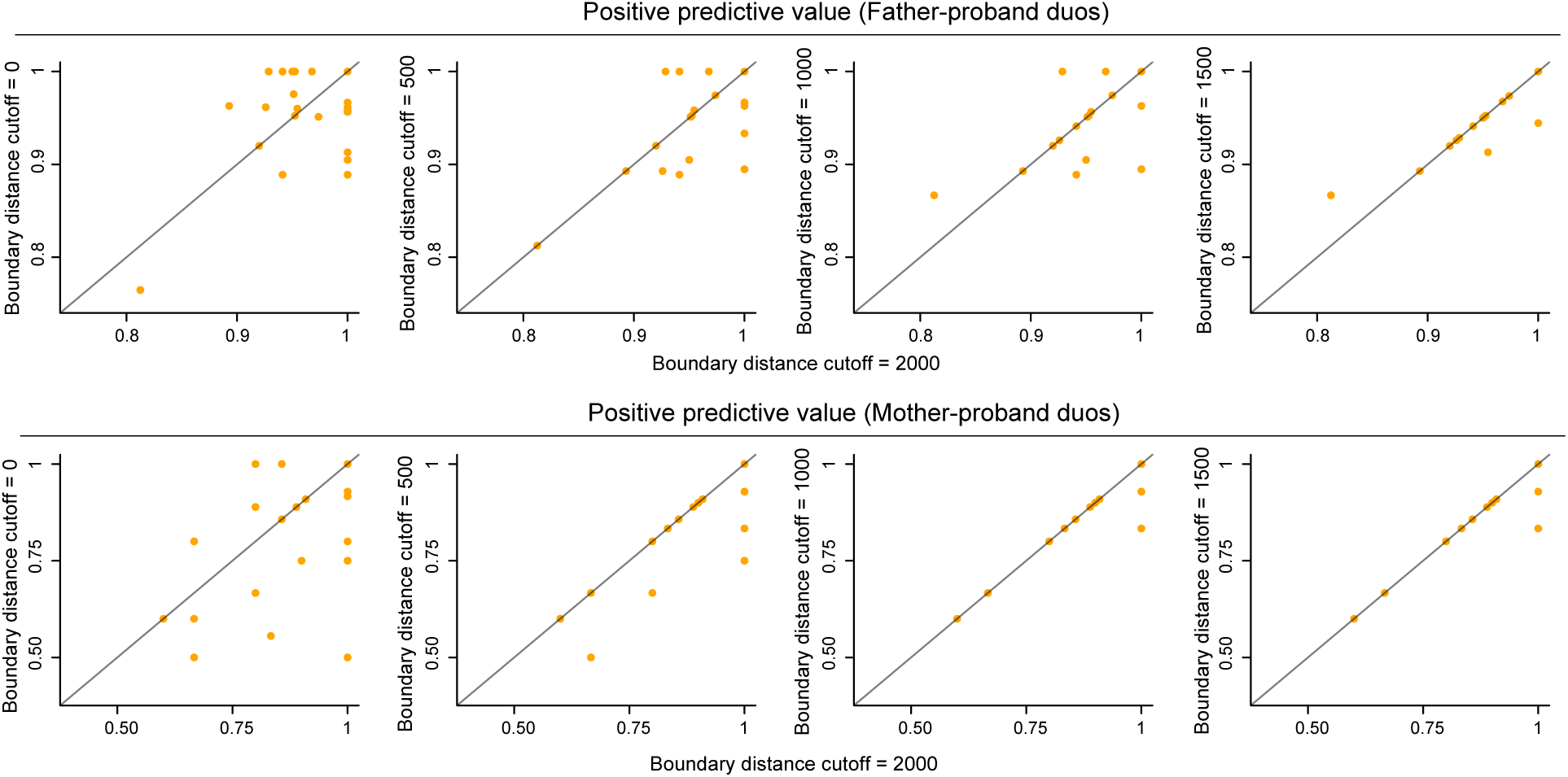
The impact of the distance threshold from haplotype block boundaries on the positive predictive value. Like Supplemental Figure S10, but showing the impact of the threshold for the distance from the haplotype block boundaries (start/end coordinates) on the PPV.

**Supplemental Figure S13.**
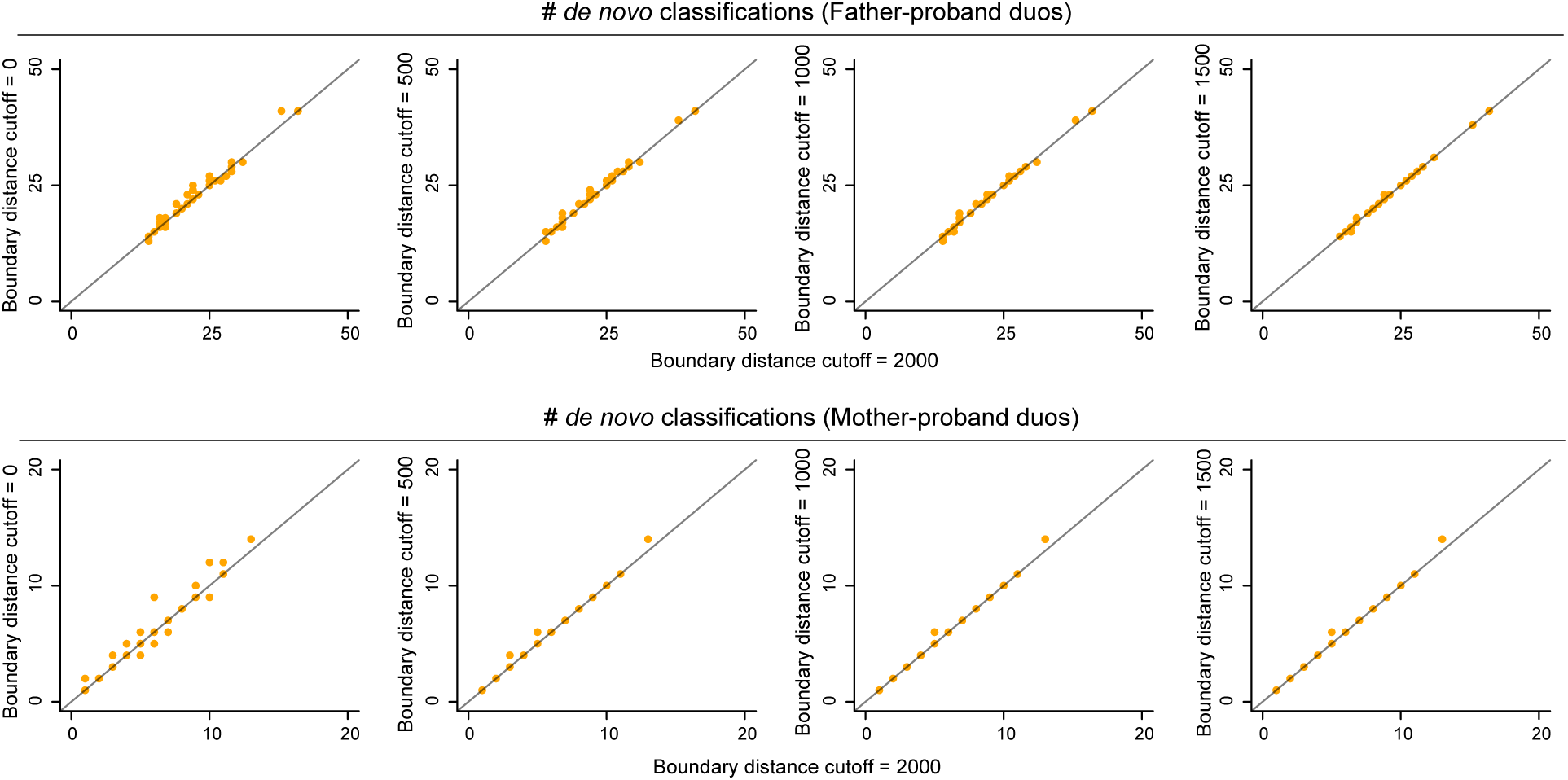
The impact of the distance threshold from haplotype block boundaries on the number of *de novo* classifications. Like Supplemental Figure S11, but showing the impact of the threshold for the distance from the haplotype block boundaries (start/end coordinates) on the number of *de novo* classifications.

**Supplemental Figure S14.**
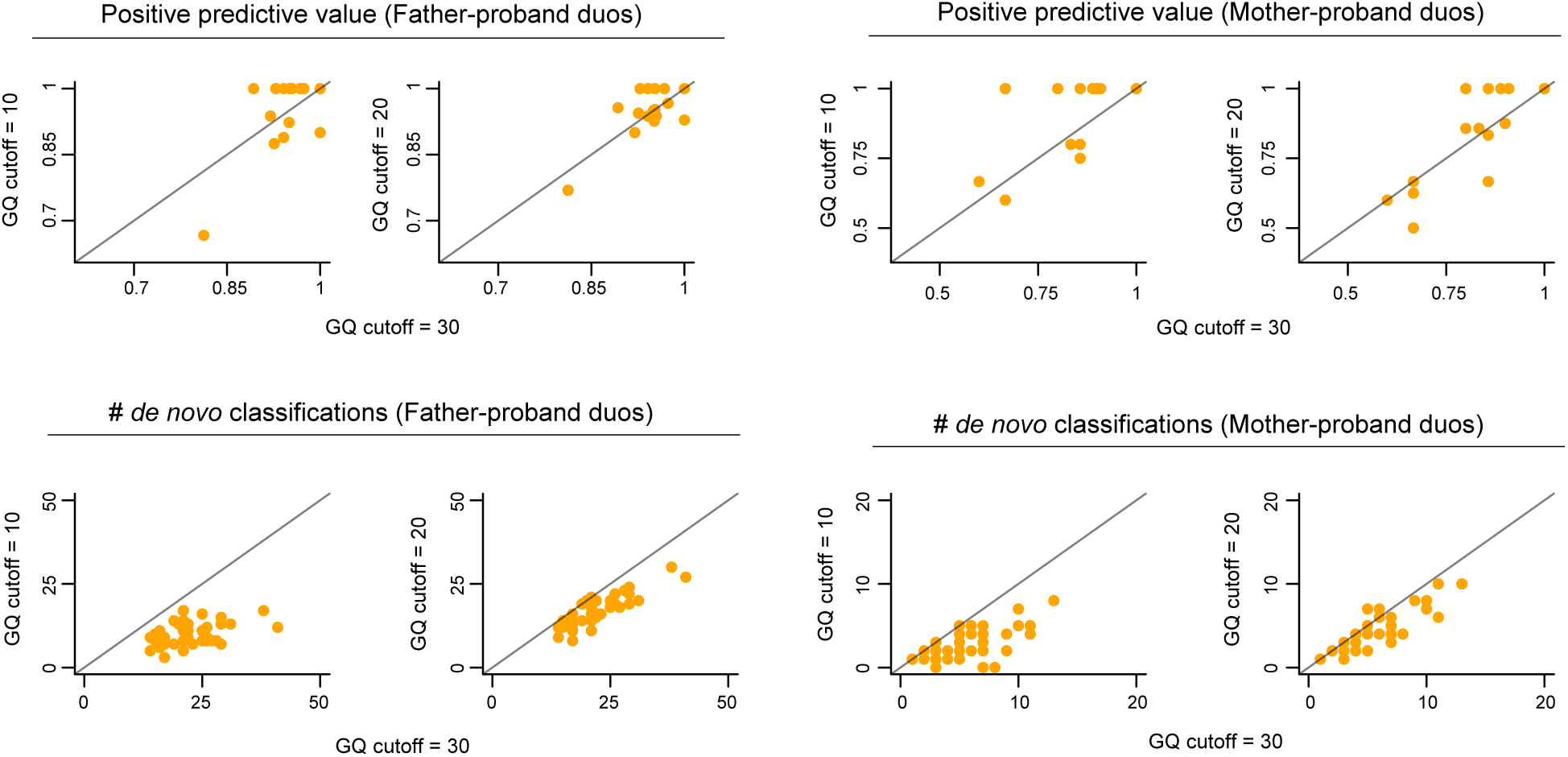
The impact of the phred quality (GQ) threshold at positions surrounding candidate variants on the positive predictive value and the number of *de novo* classifications. Like Supplemental Figure S10 and Supplemental Figure S11, but showing the impact of the GQ threshold for positions surrounding the candidate variant (that is, positions which determine the Hamming distance) on the PPV and the number of *de novo* classifications.

**Supplemental Figure S15.**
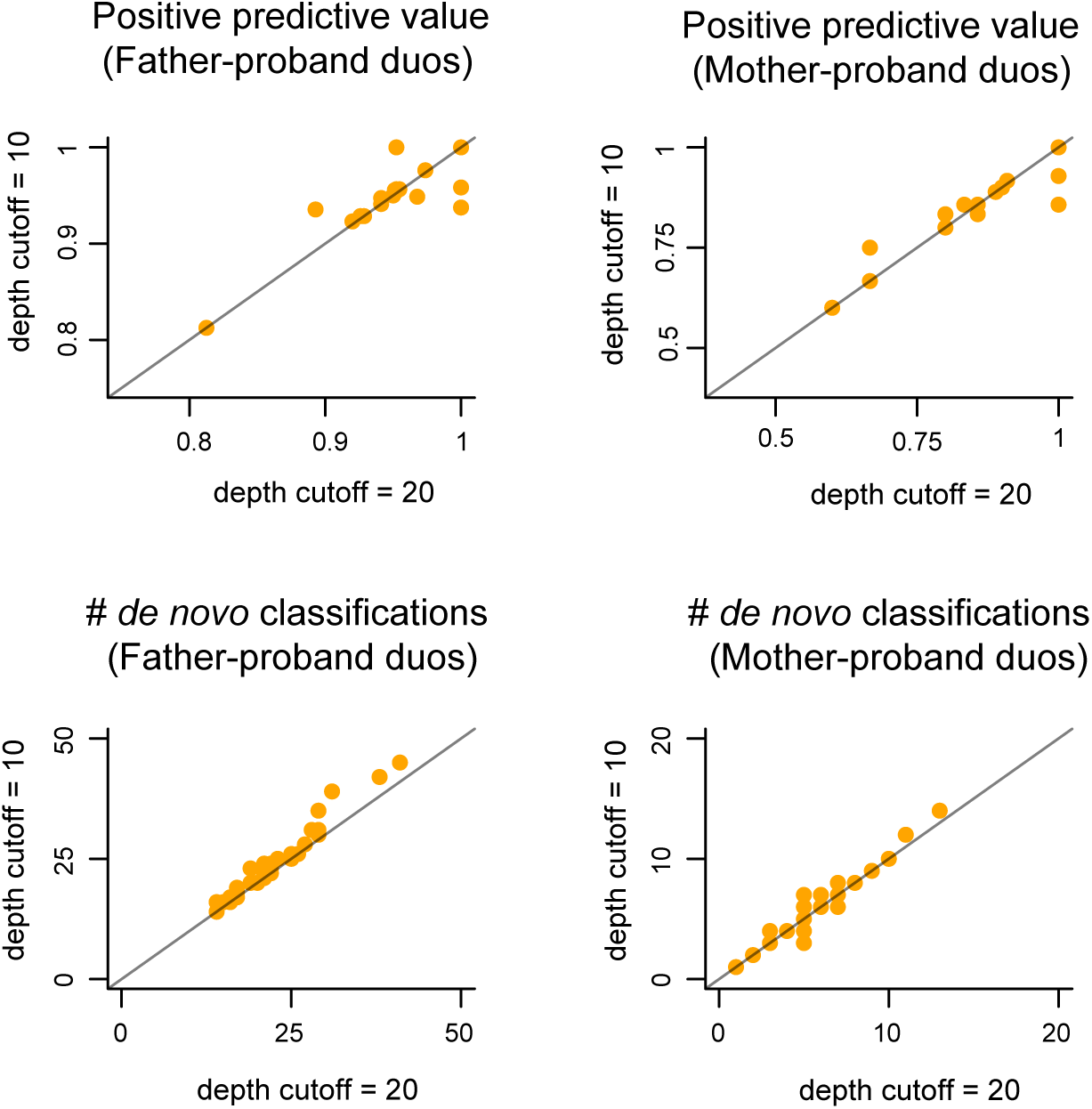
The impact of the sequencing depth threshold at positions surrounding candidate variants on the positive predictive value and the number of *de novo* classifications. Like Supplemental Figure S14, but showing the impact of the sequencing depth threshold for positions surrounding the candidate variant (that is, positions which determine the Hamming distance) on the PPV and the number of *de novo* classifications.

## References

1. Beyter D, Ingimundardottir H, Oddsson A, Eggertsson HP, Bjornsson E, Jonsson H, Atlason BA, Kristmundsdottir S, Mehringer S, Hardarson MT, et al. (2021). Long-read sequencing of 3,622 Icelanders provides insight into the role of structural variants in human diseases and other traits. Nat. Genet. 53.6: 779–786. D O I: 10.1038/s41588-021-00865-4.

2. Browning SR and Browning BL (2011). Haplotype phasing: existing methods and new developments. Nat. Rev. Genet. 12.10: 703–714. D O I: 10.1038/nrg3054.

3. Brunet T, Jech R, Brugger M, Kovacs R, Alhaddad B, Leszinski G, Riedhammer KM, Westphal DS, Mahle I, Mayerhanser K, et al. (2021). De novo variants in neurodevelopmental disorders-experiences from a tertiary care center. Clin. Genet. 100.1: 14–28. D O I: 10.1111/cge.13946.

4. Chaisson MJP, Huddleston J, Dennis MY, Sudmant PH, Malig M, Hormozdiari F, Antonacci F, Surti U, Sandstrom R, Boitano M, et al. (2015). Resolving the complexity of the human genome using single-molecule sequencing. Nature 517.7536: 608–611. D O I: 10.1038/nature13907.

5. Chen Y, Dawes R, Kim HC, Ljungdahl A, Stenton SL, Walker S, Lord J, Lemire G, Martin-Geary AC, Ganesh VS, et al. (2024). De novo variants in the RNU4-2 snRNA cause a frequent neurodevelopmental syndrome. Nature 632.8026: 832–840. D O I: 10.1038/s41586-024-07773-7.

6. Cohen ASA, Farrow EG, Abdelmoity AT, Alaimo JT, Amudhavalli SM, Anderson JT, Bansal L, Bartik L, Baybayan P, Belden B, et al. (2022). Genomic answers for children: Dynamic analyses of ¿1000 pediatric rare disease genomes. Genet. Med. 24.6: 1336–1348. D O I: 10.1016/j.gim.2022.02.007.

7. Dominguez Gonzalez CA, Bell KM, Rajagopalan R, Silva MG de, Lemes A, Zabala C, Pérez-Vidarte F, Cerisola A, Vossough A, Whitehead MT, et al. (2025). Deep intronic SVA E insertion identified as the most common pathogenic variant associated with Canavan disease: A diagnostic blind spot. Neurol. Genet. 11.5: e200291. D O I: 10.1212/NXG.0000000000200291.

8. Dwarshuis N, Kalra D, McDaniel J, Sanio P, Alvarez Jerez P, Jadhav B, Huang WE, Mondal R, Busby B, Olson ND, et al. (2024). The GIAB genomic stratifications resource for human reference genomes. Nat. Commun. 15.1: 9029. D O I: 10.1038/s41467-024-53260-y.

9. Figueroa KP, Gross C, Buena-Atienza E, Paul S, Gandelman M, Kakar N, Sturm M, Casadei N, Admard J, Park J, et al. (2024). A GGC-repeat expansion in ZFHX3 encoding polyglycine causes spinocerebellar ataxia type 4 and impairs autophagy. Nat. Genet. 56.6: 1080–1089. D O I: 10.1038/s41588-024-01719-5.

10. Greene D, Thys C, Berry IR, Jarvis J, Ortibus E, Mumford AD, Freson K, and Turro E (2024). Mutations in the U4 snRNA gene RNU4-2 cause one of the most prevalent monogenic neurodevelopmental disorders. Nat. Med. 30.8: 2165–2169. D O I: 10.1038/s41591-024-03085-5.

11. Guo MH, Francioli LC, Stenton SL, Goodrich JK, Watts NA, Singer-Berk M, Groopman E, Darnowsky PW, Solomonson M, Baxter S, et al. (2024). Inferring compound heterozygosity from large-scale exome sequencing data. Nat. Genet. 56.1: 152–161. D O I: 10.1038/s41588-023-01608-3.

12. Gustafson JA, Gibson SB, Damaraju N, Zalusky MP, Hoekzema K, Twesigomwe D, Yang L, Snead AA, Richmond PA, De Coster W, et al. (2024). High-coverage nanopore sequencing of samples from the 1000 Genomes Project to build a comprehensive catalog of human genetic variation. Genome Res.: gr.279273.124. D O I: 10.1101/gr.279273.124.

13. Hiatt SM, Lawlor JMJ, Handley LH, Latner DR, Bonnstetter ZT, Finnila CR, Thompson ML, Boston LB, Williams M, Rodriguez Nunez I, et al. (2024). Long-read genome sequencing and variant reanalysis increase diagnostic yield in neurodevelopmental disorders. Genome Res. D O I: 10.1101/gr.279227.124.

14. Holt JM, Saunders CT, Rowell WJ, Kronenberg Z, Wenger AM, and Eberle M (2024). HiPhase: jointly phasing small, structural, and tandem repeat variants from HiFi sequencing. Bioinformatics 40.2. D O I: 10.1093/bioinformatics/btae042.

15. Jaganathan K, Kyriazopoulou Panagiotopoulou S, McRae JF, Darbandi SF, Knowles D, Li YI, Kosmicki JA, Arbelaez J, Cui W, Schwartz GB, et al. (2019). Predicting splicing from primary sequence with deep learning. Cell 176.3: 535–548.e24. D O I: 10.1016/j.cell.2018.12.015.

16. Jansen S, Hoischen A, Coe BP, Carvill GL, Van Esch H, Bosch DGM, Andersen UA, Baker C, Bauters M, Bernier RA, et al. (2018). A genotype-first approach identifies an intellectual disability-overweight syndrome caused by PHIP haploinsufficiency. Eur. J. Hum. Genet. 26.1: 54–63. D O I: 10.1038/s41431-017-0039-5.

17. Jó nsson H, Sulem P, Kehr B, Kristmundsdottir S, Zink F, Hjartarson E, Hardarson MT, Hjorleifsson KE, Eggertsson HP, Gudjonsson SA, et al. (2017). Parental influence on human germline de novo mutations in 1,548 trios from Iceland. Nature 549.7673: 519–522. D O I: 10.1038/nature24018.

18. Karczewski KJ, Francioli LC, Tiao G, Cummings BB, Alföldi J, Wang Q, Collins RL, Laricchia KM, Ganna A, Birnbaum DP, et al. (2020). The mutational constraint spectrum quantified from variation in 141,456 humans. Nature 581.7809: 434–443. D O I: 10.1038/s41586-020-2308-7.

19. Kessler MD, Loesch DP, Perry JA, Heard-Costa NL, Taliun D, Cade BE, Wang H, Daya M, Ziniti J, Datta S, et al. (2020). De novo mutations across 1,465 diverse genomes reveal mutational insights and reductions in the Amish founder population. Proc. Natl. Acad. Sci. U. S. A. 117.5: 2560–2569. D O I: 10.1073/pnas.1902766117.

20. Kolmogorov M, Billingsley KJ, Mastoras M, Meredith M, Monlong J, Lorig-Roach R, Asri M, Alvarez Jerez P, Malik L, Dewan R, et al. (2023). Scalable Nanopore sequencing of human genomes provides a comprehensive view of haplotype-resolved variation and methylation. Nat. Methods 20.10: 1483–1492. D O I: 10.1038/s41592-023-01993-x.

21. Kong A, Frigge ML, Masson G, Besenbacher S, Sulem P, Magnusson G, Gudjonsson SA, Sigurdsson A, Jonasdottir A, Jonasdottir A, et al. (2012). Rate of de novo mutations and the importance of father’s age to disease risk. Nature 488.7412: 471–475. D O I: 10.1038/nature11396.

22. Kucuk E, Sanden BPGH van der, O’Gorman L, Kwint M, Derks R, Wenger AM, Lambert C, Chakraborty S, Baybayan P, Rowell WJ, et al. (2023). Comprehensive de novo mutation discovery with HiFi long-read sequencing. Genome Med. 15.1: 34. D O I: 10.1186/s13073-023-01183-6.

23. Ligt J de, Willemsen MH, Bon BWM van, Kleefstra T, Yntema HG, Kroes T, Vulto-van Silfhout AT, Koolen DA, Vries P de, Gilissen C, et al. (2012). Diagnostic exome sequencing in persons with severe intellectual disability. N. Engl. J. Med. 367.20: 1921–1929. D O I: 10.1056/NEJMoa1206524.

24. Logsdon GA, Vollger MR, and Eichler EE (2020). Long-read human genome sequencing and its applications. Nat. Rev. Genet. 21.10: 597–614. D O I: 10.1038/s41576-020-0236-x.

25. Manders F, Brandsma AM, Kanter J de, Verheul M, Oka R, Roosmalen MJ van, Roest B van der, Hoeck A van, Cuppen E, and Boxtel R van (2022). MutationalPatterns: the one stop shop for the analysis of mutational processes. BMC Genomics 23.1: 134. D O I: 10.1186/s12864-022-08357-3.

26. Martin M, Patterson M, Garg S, O Fischer S, Pisanti N, Klau GW, Schöenhuth A, and Marschall T (2016). WhatsHap: fast and accurate read-based phasing. bioRxiv. D O I: 10.1101/085050.

27. Mastrorosa FK, Miller DE, and Eichler EE (2023). Applications of long-read sequencing to Mendelian genetics. Genome Med. 15.1: 42. D O I: 10.1186/s13073-023-01194-3.

28. McCombie WR and McPherson JD (2019). Future promises and concerns of ubiquitous next-generation sequencing. Cold Spring Harb. Perspect. Med. 9.9: a025783. D O I: 10.1101/cshperspect.a025783.

29. Merker JD, Wenger AM, Sneddon T, Grove M, Zappala Z, Fresard L, Waggott D, Utiramerur S, Hou Y, Smith KS, et al. (2018). Long-read genome sequencing identifies causal structural variation in a Mendelian disease. Genet. Med. 20.1: 159–163. D O I: 10.1038/gim.2017.86.

30. Miller DE, Sulovari A, Wang T, Loucks H, Hoekzema K, Munson KM, Lewis AP, Fuerte EPA, Paschal CR, Walsh T, et al. (2021). Targeted long-read sequencing identifies missing disease-causing variation. Am. J. Hum. Genet. 108.8: 1436–1449. D O I: 10.1016/j.ajhg.2021.06.006.

31. Mizuguchi T, Okamoto N, Yanagihara K, Miyatake S, Uchiyama Y, Tsuchida N, Hamanaka K, Fujita A, Miyake N, and Matsumoto N (2021). Pathogenic 12-kb copy-neutral inversion in syndromic intellectual disability identified by high-fidelity long-read sequencing. Genomics 113.1 Pt 2: 1044–1053. D O I: 10.1016/j.ygeno.2020.10.038.

32. Negi S, Stenton SL, Berger SI, Canigiula P, McNulty B, Violich I, Gardner J, Hillaker T, O’Rourke SM, O’Leary MC, et al. (2025). Advancing long-read nanopore genome assembly and accurate variant calling for rare disease detection. Am. J. Hum. Genet. D O I: 10.1016/j.ajhg.2025. 01.002.

33. Obenchain V, Lawrence M, Carey V, Gogarten S, Shannon P, and Morgan M (2014). VariantAnnotation: a Bioconductor package for exploration and annotation of genetic variants. Bioinformatics 30.14: 2076–2078. D O I: 10.1093/bioinformatics/btu168.

34. Pedersen BS, Bhetariya PJ, Brown J, Kravitz SN, Marth G, Jensen RL, Bronner MP, Underhill HR, and Quinlan AR (2020). Somalier: rapid relatedness estimation for cancer and germline studies using efficient genome sketches. Genome Med. 12.1: 62. D O I: 10.1186/s13073-020-00761-2.

35. Poplin R, Chang PC, Alexander D, Schwartz S, Colthurst T, Ku A, Newburger D, Dijamco J, Nguyen N, Afshar PT, et al. (2018). A universal SNP and small-indel variant caller using deep neural networks. Nat. Biotechnol. 36.10: 983–987. D O I: 10.1038/nbt.4235.

36. Posey JE, Harel T, Liu P, Rosenfeld JA, James RA, Coban Akdemir ZH, Walkiewicz M, Bi W, Xiao R, Ding Y, et al. (2017). Resolution of disease phenotypes resulting from multilocus genomic variation. N. Engl. J. Med. 376.1: 21–31. D O I: 10.1056/NEJMoa1516767.

37. Rehm HL, Alaimo JT, Aradhya S, Bayrak-Toydemir P, Best H, Brandon R, Buchan JG, Chao EC, Chen E, Clifford J, et al. (2023). The landscape of reported VUS in multi-gene panel and genomic testing: Time for a change. Genet. Med. 25.12: 100947. D O I: 10.1016/j.gim.2023.100947.

38. Richards S, Aziz N, Bale S, Bick D, Das S, Gastier-Foster J, Grody WW, Hegde M, Lyon E, Spector E, et al. (2015). Standards and guidelines for the interpretation of sequence variants: a joint consensus recommendation of the American College of Medical Genetics and Genomics and the Association for Molecular Pathology. Genet. Med. 17.5: 405–424. D O I: 10.1038/gim.2015.30.

39. Sasani TA, Pedersen BS, Gao Z, Baird L, Przeworski M, Jorde LB, and Quinlan AR (2019). Large, three-generation human families reveal post-zygotic mosaicism and variability in germline mutation accumulation. Elife 8. D O I: 10.7554/eLife.46922.

40. Sedlazeck FJ, Rescheneder P, Smolka M, Fang H, Nattestad M, Haeseler A von, and Schatz MC (2018). Accurate detection of complex structural variations using single-molecule sequencing. Nat. Methods 15.6: 461–468. D O I: 10.1038/s41592-018-0001-7.

41. Shojaeisaadi H, Schoenrock A, Meier MJ, Williams A, Norris JM, Palmer ND, Yauk CL, and Marchetti F (2024). Mutational signature analyses in multi-child families reveal sources of age-related increases in human germline mutations. *Commun*. Biol. 7.1: 1451. 10.1038/s42003-024-07140-2.

42. Smolka M, Paulin LF, Grochowski CM, Horner DW, Mahmoud M, Behera S, Kalef-Ezra E, Gandhi M, Hong K, Pehlivan D, et al. (2024). Detection of mosaic and population-level structural variants with Sniffles2. Nat. Biotechnol. 42.10: 1571–1580. 10.1038/s41587-023-02024-y.

43. Steyaert W, Sagath L, Demidov G, Yépez VA, Esteve-Codina A, Gagneur J, Ellwanger K, Derks R, Weiss M, Ouden A den, et al. (2024). Unravelling undiagnosed rare disease cases by HiFi long-read genome sequencing. *medRxiv*. 10.1101/2024.05.03.24305331.

44. Vissers LELM, Ligt J de, Gilissen C, Janssen I, Steehouwer M, Vries P de, Lier B van, Arts P, Wieskamp N, Rosario M del, et al. (2010). A de novo paradigm for mental retardation. Nat. Genet. 42.12: 1109–1112. D O I: 10.1038/ng.712.

45. Vollger MR, Korlach J, Eldred KC, Swanson E, Underwood JG, Bohaczuk SC, Mao Y, Cheng YHH, Ranchalis J, Blue EE, et al. (2025). Synchronized long-read genome, methylome, epigenome and transcriptome profiling resolve a Mendelian condition. Nat. Genet. D O I: 10.1038/s41588-024-02067-0.

46. Walker LC, Hoya Mdl, Wiggins GAR, Lindy A, Vincent LM, Parsons MT, Canson DM, Bis-Brewer D, Cass A, Tchourbanov A, et al. (2023). Using the ACMG/AMP framework to capture evidence related to predicted and observed impact on splicing: Recommendations from the ClinGen SVI Splicing Subgroup. Am. J. Hum. Genet. 110.7: 1046–1067. 10.1016/j.ajhg.2023.06.002.

47. Wang K, Li M, and Hakonarson H (2010). ANNOVAR: functional annotation of genetic variants from high-throughput sequencing data. Nucleic Acids Res. 38.16: e164. 10.1093/nar/gkq603.

48. Webster E, Cho MT, Alexander N, Desai S, Naidu S, Bekheirnia MR, Lewis A, Retterer K, Juusola J, and Chung WK (2016). De novo PHIP-predicted deleterious variants are associated with developmental delay, intellectual disability, obesity, and dysmorphic features. Cold Spring Harb. Mol. Case Stud. 2.6: a001172. 10.1101/mcs.a00117.

49. Wenger AM, Peluso P, Rowell WJ, Chang PC, Hall RJ, Concepcion GT, Ebler J, Fungtammasan A, Kolesnikov A, Olson ND, et al. (2019). Accurate circular consensus long-read sequencing improves variant detection and assembly of a human genome. Nat. Biotechnol. 37.10: 1155–1162. D O I: 10.1038/s41587-019-0217-9.

50. Wojcik MH, Lemire G, Berger E, Zaki MS, Wissmann M, Win W, White SM, Weisburd B, Wieczorek D, Waddell LB, et al. (2024). Genome sequencing for diagnosing rare diseases. N. Engl. J. Med. 390.21: 1985–1997. D O I: 10.1056/NEJMoa2314761.

51. Wojcik MH, Reuter CM, Marwaha S, Mahmoud M, Duyzend MH, Barseghyan H, Yuan B, Boone PM, Groopman EE, Délot EC, et al. (2023). Beyond the exome: What’s next in diagnostic testing for Mendelian conditions. Am. J. Hum. Genet. 110.8: 1229–1248. 10.1016/j.ajhg.2023.06.009.

52. Wright CF, Campbell P, Eberhardt RY, Aitken S, Perrett D, Brent S, Danecek P, Gardner EJ, Chundru VK, Lindsay SJ, et al. (2023). Genomic diagnosis of rare pediatric disease in the United Kingdom and Ireland. N. Engl. J. Med. 388.17: 1559–1571. D O I: 10.1056/NEJMoa2209046.

53. Xie Z, Sun C, Zhang S, Liu Y, Yu M, Zheng Y, Meng L, Acharya A, Cornejo-Sanchez DM, Wang G, et al. (2020). Long-read whole-genome sequencing for the genetic diagnosis of dystrophinopathies. Ann. Clin. Transl. Neurol. 7.10: 2041–2046. D O I: 10.1002/acn3.51201.

54. Yang Y, Muzny DM, Xia F, Niu Z, Person R, Ding Y, Ward P, Braxton A, Wang M, Buhay C, et al. (2014). Molecular findings among patients referred for clinical whole-exome sequencing. JAMA 312.18: 1870. D O I: 10.1001/jama.2014.14601.

55. Yun T, Li H, Chang PC, Lin MF, Carroll A, and McLean CY (2021). Accurate, scalable cohort variant calls using DeepVariant and GLnexus. Bioinformatics 36.24: 5582–5589. 10.1093/bioinformatics/btaa1081.

